# Wrong person, place and time: viral load and contact network structure predict SARS-CoV-2 transmission and super-spreading events

**DOI:** 10.1101/2020.08.07.20169920

**Authors:** Ashish Goyal, Daniel B. Reeves, E. Fabian Cardozo-Ojeda, Joshua T. Schiffer, Bryan T. Mayer

## Abstract

SARS-CoV-2 is difficult to contain because many transmissions occur during the pre-symptomatic phase of infection. Moreover, in contrast to influenza, while most SARS-CoV-2 infected people do not transmit the virus to anybody, a small percentage secondarily infect large numbers of people. We designed mathematical models of SARS-CoV-2 and influenza which link observed viral shedding patterns with key epidemiologic features of each virus, including distributions of the number of secondary cases attributed to each infected person (individual R_0_) and the duration between symptom onset in the transmitter and secondarily infected person (serial interval). We identify that people with SARS-CoV-2 or influenza infections are usually contagious for fewer than one day congruent with peak viral load several days after infection, and that transmission is unlikely below a certain viral load. SARS-CoV-2 super-spreader events with over 10 secondary infections occur when an infected person is briefly shedding at a very high viral load and has a high concurrent number of exposed contacts. The higher predisposition of SARS-CoV-2 towards super-spreading events is not due to its 1-2 additional weeks of viral shedding relative to influenza. Rather, a person infected with SARS-CoV-2 exposes more people within equivalent physical contact networks than a person infected with influenza, likely due to aerosolization of virus. Our results support policies that limit crowd size in indoor spaces and provide viral load benchmarks for infection control and therapeutic interventions intended to prevent secondary transmission.

**One Sentence Summary:** We developed a coupled within-host and between-host mathematical model to identify viral shedding levels required for transmission of SARS-CoV-2 and influenza, and to explain why super-spreading events occur more commonly during SARS-CoV-2 infection.

## Introduction

The SARS-CoV-2 pandemic is an ongoing tragedy that has caused 700,000 deaths and massively disrupted the global economy. The pandemic is rapidly expanding in the United States and is re-emerging focally in many countries that had previous success in limiting its spread.^1^

Two features have proven challenging in containing outbreaks. First, most transmissions occur during the pre-symptomatic phase of infection.^2^ Underlying this observation is a highly variable incubation period, defined as time between infection and symptom onset, which often extends beyond an infected person’s peak viral shedding.^3^

Second, there is substantial over-dispersion of the basic reproduction number (R0) for an individual infected with SARS-CoV-2,^4^ meaning that most infected people do not transmit at all, while a minority may transmit to dozens of people, with the average, population R0 achieving a high enough level (>1) to allow exponential growth of cases in the absence of an effective intervention.^5^ Approximately 10-20% of infected people account for 80% of SARS-CoV-2 transmissions.^4,6^ Super-spreader events, in which the duration of contact between a single transmitter and large number of secondarily infected people is often limited to hours, are well documented.^7,8^ This pattern is not evident for influenza which has more homogeneous individual transmissions numbers.^9,10^ Differing shedding kinetics between the two viruses might explain this distinction; SARS-CoV-2 is often present intermittently in the upper airways for many weeks,^11,12^ while influenza is rarely shed for more than a week.^13^ Alternatively, SARS-CoV-2 aerosolization may predispose to wider exposure networks given the presence of an infected person in a crowded indoor space.

Viral load is recognized as a strong determinant of transmission risk. For influenza, the dose of viral exposure is related to the probability of infection in human challenge studies,^14^ and early treatment reduces household transmission.^15,16^ Household shedding of human herpesvirus-6 is closely linked to subsequent infection in newborns,^17^ and infants shedding high levels of cytomegalovirus in the oropharynx predictably transmit the virus back to their mothers.^18^

The epidemiology of viral infections can also be perturbed by biomedical interventions that lower viral load at mucosal transmission surfaces. Reduction of genital herpes simplex virus-2 shedding with antiviral treatments decreases probability of transmission.^19^ Suppressive antiretroviral therapy (ART) for HIV virtually eliminates the possibility of partner-to-partner sexual transmission and has limited community transmission dramatically.^20,21^

These concepts are relevant for SARS-CoV-2 infection and require urgent attention as the pandemic continues to wreak havoc. Early therapies that lower peak viral load may reduce the severity of COVID-19 but may also decrease the probability of transmission and of super-spreader events.^22^ Similarly, the effectiveness of policies such as limiting mass gatherings, and enforcing mask use can be directly evaluated by their ability to reduce exposure viral load and transmission risk.^23^ Here we developed a transmission simulation framework to capture the contribution of viral load to observed epidemiologic transmission metrics for influenza and SARS-CoV-2 and used this approach to explain why SARS-CoV-2 is predisposed to super-spreading events.

## Results

### Overall approach

We designed a series of steps to estimate the viral load required for SARS-CoV-2 and influenza transmission, as well as conditions required to explain the observed over-dispersion of secondary infections *(individual R0)* and frequent super-spreader events associated with SARS-CoV-2 but not influenza. This process included within-host modeling of viral loads, simulations of exposures and possible transmissions based on various transmission dose response curves, testing of various parameter sets against epidemiologic data and exploratory analyses with the best fitting model (**Fig S1**).

### Within-host mathematical model of SARS CoV-2 shedding

First, we used our previously developed within-host mathematical model (equations in the **Methods**),^24^ to generate plausible viral load patterns in the upper airway of an infected person or *transmitter* who could potentially transmit the virus to others (**Fig 1, Fig S2a**). Briefly, the model captures observed upper airway viral kinetics from 25 people from four different countries.^25-28^ Key observed features include an early viral peak followed by a decelerating viral clearance phase, which in turn leads to a temporary plateau at a lower viral load, ultimately followed by rapid viral elimination. Our model captures these patterns by including a density dependent term for early infected cell elimination and a nonspecific acquired immune term for late infected cell elimination.

**Fig 1.**
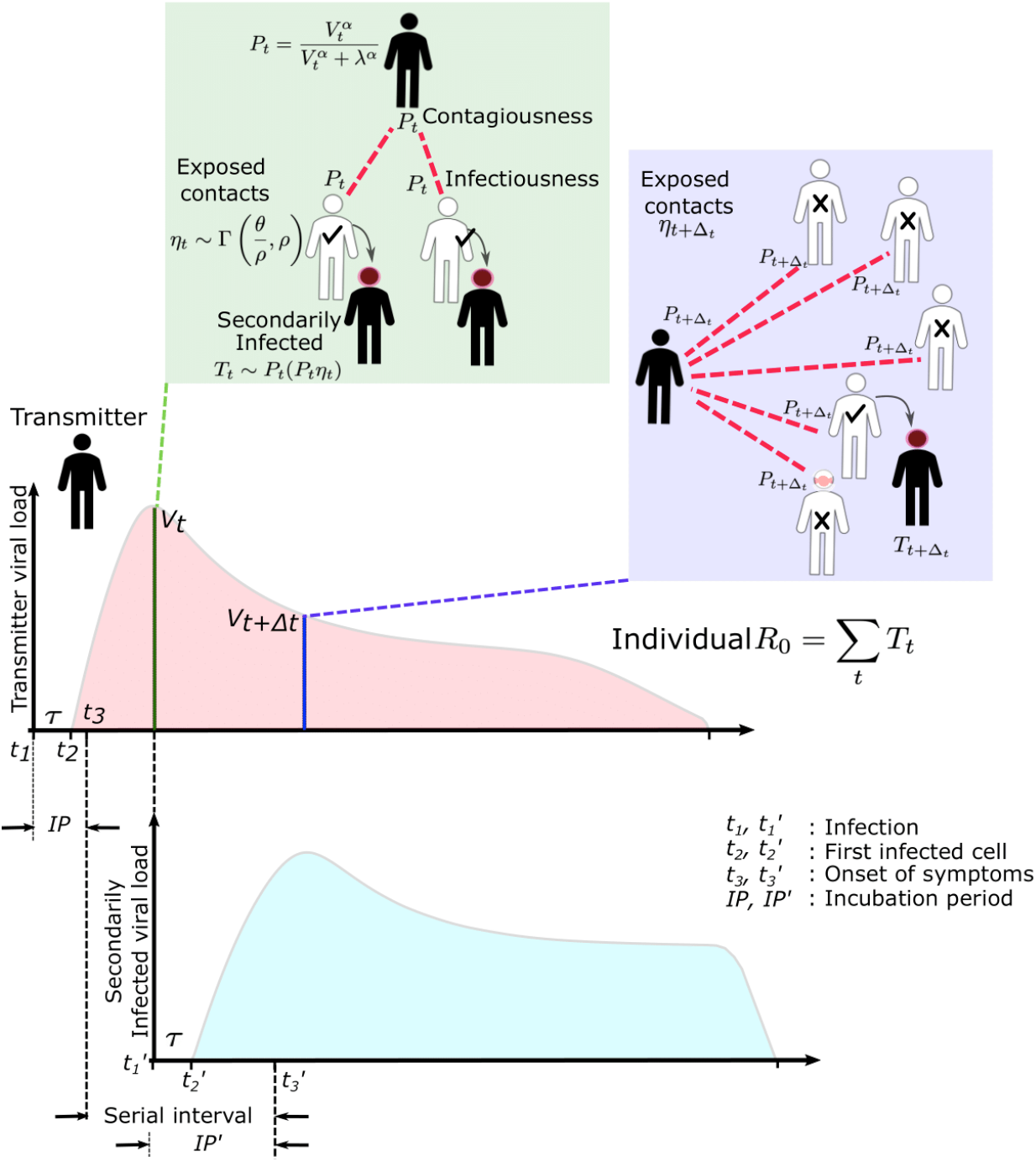
SARS-CoV-2 and influenza transmission model schematic. In the above cartoon, the transmitter has 2 exposure events at discrete timepoints resulting in 7 total exposure contacts and 3 secondary infections. Transmission is more likely at the first exposure event due to higher exposure viral load. To model this process, the timing of exposure events and number of exposed contacts is governed by a random draw from a gamma distribution which allows for heterogeneity in number of exposed contacts per day (**Fig S3**). Viral load is sampled at the precise time of each exposure event. Probability of transmission is identified based on the product of two dose curves (**Fig S2C, D**) which capture contagiousness (probability of viral passage to an exposure contact’s airway) and infectiousness (probability of transmission given viral presence in the airway). Incubation period (**Fig S4**) of the transmitter and secondarily infected person is an input into each simulation and is depicted graphically. Individual R0 is an output of each simulation and is defined as the number of secondary infections generated by an infected individual. Serial interval is an output of each simulated transmission and is depicted graphically.

One limitation of our model is that only half of study participants provided longitudinal viral load data from the very early days of infection when COVID-19 is often pre-symptomatic. Therefore, the model’s output is most reliable for later time points. In particular, we have somewhat limited information on viral expansion rate and duration of peak shedding. To impute possible variability, we generated a set of heterogeneous shedding curves in which the viral upslope, the downslope of viral load after peak and the viral load during plateau phase were varied (**Fig S2b**). Overall, the model generated several distinct patterns of infection: rapid elimination after the initial peak, a prolonged plateau phase with a low viral load, and a prolonged plateau phase with higher viral load. We simulated the transmission model with and without imputed heterogeneity.

### Transmission dose response curves

We defined an *exposure event* in very specific biologic terms as a discrete event consisting of sufficient contact in time and space between a transmitter and one or more uninfected persons *(exposure contacts)* to allow for the possibility of a successful transmission. We next designed hundreds of dose response curves which separately predict contagiousness (CD curves) and infectiousness (ID curves) at a certain viral dose given an exposure contact. *Contagiousness* is defined as the viral load dependent probability of passage of virus-laden droplets or airborne particles from the airways of a potential transmitter to the airway of an exposure contact. *Infectiousness* is defined as the viral load dependent probability of transmission given direct airway exposure to virus in an exposure contact. *Transmission risk* is the product of these two mechanistic probabilities derived from the ID and CD curves and results is a transmission dose (TD) response curve. Each CD or ID curve is defined by its ID50 (*λ*) or viral load at which contagion or infection probability is 50% (**Fig S2c**), as well as its slope (*α*) (**Fig S2d**).^29^ The TD50 is defined as viral load at which there is 50% transmission probability. We assumed equivalent curves for contagiousness and infectiousness for model fitting purposes. We also considered a simpler model with only a single TD curve (for *infectiousness*) and obtained qualitatively similar results (**Supplement and Methods**). Our model is inclusive of the hypothesis that viral load is not a key determinant of transmission when *α*<<1 (**Fig S2d**).

### Exposure contact rate simulations

We introduced heterogeneity of exposure contact rates among possible transmitters by randomly selecting from a gamma distribution defined by mean number of exposure contacts per day (*θ*) and a scaling factor (*ρ*) that controls daily variability (**Fig S3**).

### Transmission simulations

For each defined exposure contact, viral load in the transmitter was sampled and transmission risk was then identified based on the product of the CD and ID curves, or the TD curve (**Fig S2e, f; Fig 1**). Based on these probabilities, we stochastically modeled whether a transmission occurred for each exposure contact. This process was repeated when there were multiple possible exposure events within a given discretized time interval and the total number of exposures and transmissions within that interval was calculated.

**Fig 2.**
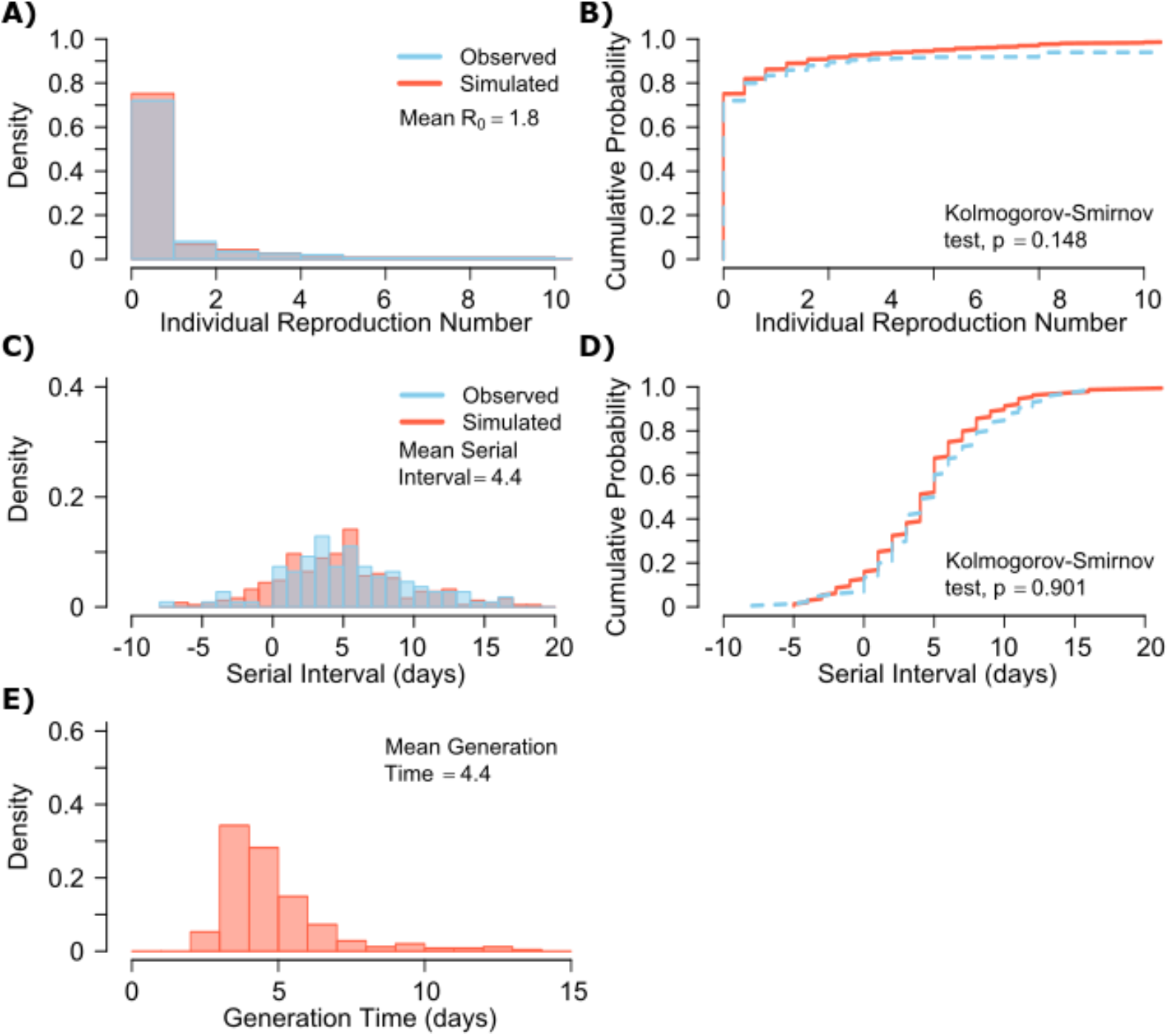
SARS-CoV-2 transmission model fit. **A**. Simulated and actual frequency histograms of individual R0 values, **B**. Simulated and actual cumulative distribution of individual R0 values. **C**. Simulated and actual frequency histograms of individual serial intervals, **D**. Simulated and actual cumulative distribution of individual serial intervals. **E**. Frequency distribution of simulated generation times.

For each successful transmission, we assumed that it takes τ days for the first infected cell to produce virus. To inform simulated values of *serial interval* (SI or time between symptom onset in the secondarily infected and transmitter), we randomly selected the *incubation period* (IP), for both the transmitter and the newly infected person, from a gamma distribution based on existing data (**Fig S4a**).^3,30^ Incubation period was defined as time from infection to the time of the onset of symptoms, where the mean incubation for SARS-CoV-2 is 5.2 days compared to 2 days for influenza.^3,9,30^

### Model fitting

In order to identify the parameter set that best recapitulated the observed data, we then simulated several hundred thousands of parameter sets with ~250 possible TD curves defined by ID50 and CD50 (*λ*) and slope (*α*), along with ~180 combinations of the mean exposed contact rate per day (*θ*) and associated variance parameter (*ρ*), and values of *τ* ∊ [0.5,1,2, 3] days. We aimed to identify the parameter set that best recapitulated the following features of the observed epidemiologic and individual-level data for SARS-CoV-2: mean R0 across individuals (R0 ∊ [1.4,2.5]),^3,4,6,31,32^ mean serial interval across individuals (SI ∊ [4.0,4.5]),^3,31,33^ cumulative distribution functions of individual R0,^4,6,34-36^ and cumulative distribution functions of serial intervals derived from SARS-CoV-2 transmission pair studies that were conducted early during the pandemic,^31^ prior to any confounding influence of social distancing measures. Here, we define *individual R0* as the total number of secondary transmissions from the transmitter in a fully susceptible population (**Methods**). Given that viral RNA is composed mostly of non-infectious material, we further checked the closeness of the solved ID curve with the observed relationship between viral RNA and probability of positive viral culture from a longitudinal cohort of infected people.^37^

### Influenza modeling

Next, we performed equivalent analyses for influenza to explain the lower frequency of observed super-spreader events with this infection. Influenza viral kinetics were modelled using a previously data-validated model.^38^ Incubation periods for influenza are lower and less variable than for SARS-CoV-2 and were randomly selected for each simulation of the model using a gamma distribution (**Fig S4b**).^39^ We again fit the model to: mean R0 across individuals (R0 ∊ [1.1,1.5]),^40-42^ mean serial interval (SI ∊ [2.9,4.3]),^9^ cumulative distribution functions of individual R0 corresponding to the 2008-2009 influenza A H1N1 pandemic with mean R0=1.26 and dispersion parameter=2.36 in the negative binomial distribution, and cumulative distribution functions of serial intervals.^9,10,40^

### Model-predicted individual R0 and serial intervals for SARS-CoV-2 infection

A single model parameter set ([*α*, *λ*, *τ*, *θ*, *ρ*] = [0.8, 10^7^, 0.5, 4, 40]) most closely reproduced empirically observed individual R0 and serial interval histograms (**Fig 2a, c**) and cumulative distribution functions (**Fig 2b, d**). We re-ran the model to fit to a higher population R0 of 2.8 and arrived at a similar set of parameter values but with a higher daily rate of exposure contacts *([α*, *λ*, *τ*, *θ*, *ρ]* = [0.8, 10^7.5^, 0.5, 20, 30]). Despite assuming that each infected person sheds at a high viral load for a period of time (**Fig 1, Fig S2b**), the model captured the fact that ~75% of 10,000 simulated transmitters do not infect any other people and that each increase in the number of possible transmissions is associated with a decreasing probability (**Fig. 2a**).

SARS-CoV-2 viral load was recently measured with viral RNA levels and mapped to concurrent probability of positive viral culture in a Dutch cohort.^37^ Our model output demonstrated a nearly equivalent infectious dose response curve if we multiplied modeled viral RNA levels by 25 (**Fig S5**): this adjustment was likely necessary because viral loads in the Dutch study participants were notably higher than those in German, Singaporean, Korean and French participants used in our intra-host model fitting.^25-28,37^

The model also generated super-spreader events with 10,000 simulated transmissions (**Fig. 2b**). If super-spreaders are defined as those who produce at least 5 secondary infections, we estimate that ~10% of all infected people and ~35% of all transmitters are super-spreaders. If super-spreaders are defined as those who produce at least 10 secondary infections, we estimate that ~6% of all infected people and ~25% of all transmitters are super-spreaders. If super-spreaders are defined as those who produce at least 20 secondary infections, we estimate that ~2.5% of all infected people and ~10% of all transmitters are super-spreaders. If super-spreaders are defined as those producing ≥5, ≥10, or ≥20 secondary infections, the contribution to all secondary infections is estimated at ~85%, ~70%, or ~44%, respectively (**Table 1**).

**Table 1:**
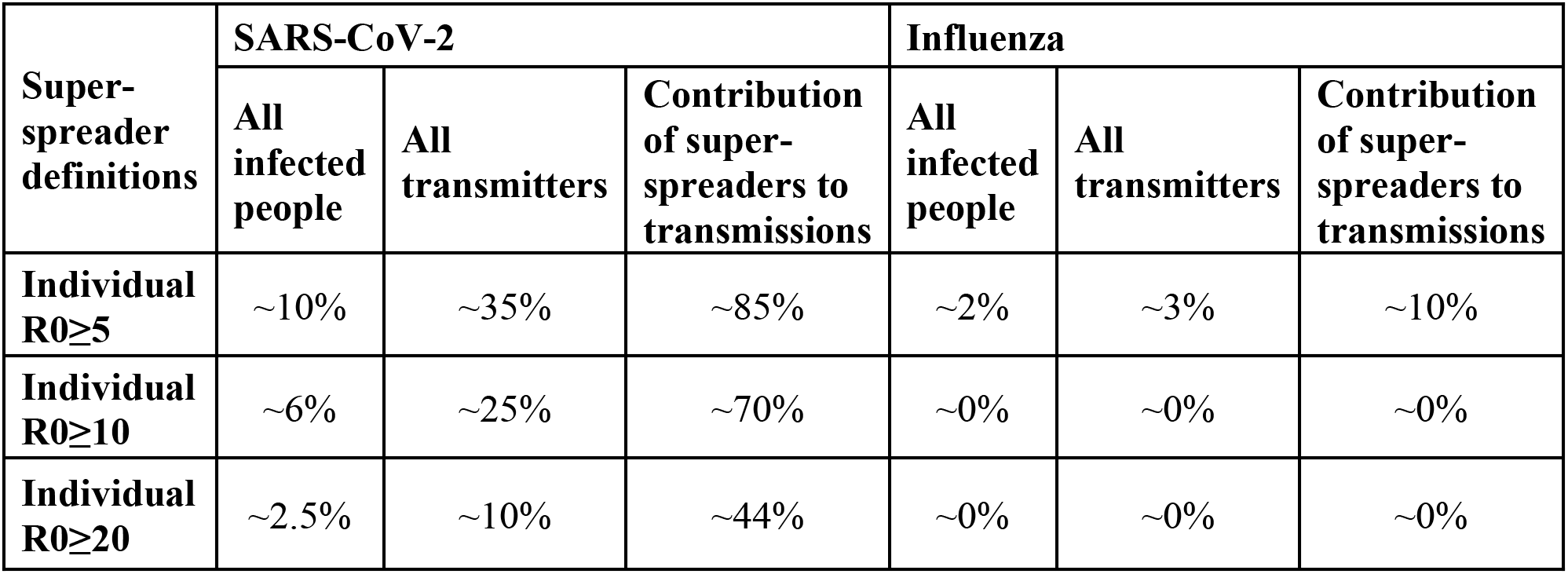
Prevalence of super-spreaders among transmitters, and contribution of super-spreading events to all SARS-CoV-2 and influenza transmissions. Estimates are from 10,000 simulations.

| Super-spreader definitions | SARS-CoV-2 |  |  | Influenza |  |  |
| --- | --- | --- | --- | --- | --- | --- |
|  | All infected people | All transmitters | Contribution of super-spreaders to transmissions | All infected people | All transmitters | Contribution of super-spreaders to transmissions |
| Individual $R_0 \geq 5$ | ~10% | ~35% | ~85% | ~2% | ~3% | ~10% |
| Individual $R_0 \geq 10$ | ~6% | ~25% | ~70% | ~0% | ~0% | ~0% |
| Individual $R_0 \geq 20$ | ~2.5% | ~10% | ~44% | ~0% | ~0% | ~0% |

The model also recapitulated the high variance of the serial interval observed within SARS-CoV-2 transmission pairs, including negative values observed in the data (**Fig 2c, d**). We next projected *generation time*, defined as the period between when an individual becomes infected and when they transmit the virus, for all transmission pairs and identified that the mean serial interval (4.4 days) provides an accurate approximation of mean generation time. However, the variance of generation time was considerably lower and by definition does not include negative values. A majority of generation times fell between 4 and 7 days, compared to −5 to 12 days for the serial interval (**Fig 2e**).

### Viral load thresholds for SARS-CoV-2 transmission

The optimized ID curve has an ID50 of 10^7^ viral RNA copies and a moderately steep slope (**Fig 3a**). The TD50 for SARS-CoV-2 was slightly higher at 10^7.5^ viral RNA copies (**Fig 3a**). To assess the impact of these parameters on transmission, we performed simulations with 10,000 transmitters and concluded that transmission is very unlikely (~0.00005%) given an exposure to an infected person with an upper airway viral load of <10^4^ SARS-CoV-2 RNA copies, and unlikely (~0.002%) given an exposure to an infected person with a viral load of <10^5^ SARS-CoV-2 RNA copies. On the other hand, transmission is much more likely (39%) given an exposure to an infected person who is shedding >10^7^ SARS-CoV-2 RNA copies, and 75% given an exposure to an infected person with a viral load of >10^8^ SARS-CoV-2 RNA copies. We obtain similar results (not shown) when we solve our model using the assumption of homogeneous viral load trajectories as in **Fig S2a**.

**Fig 3.**
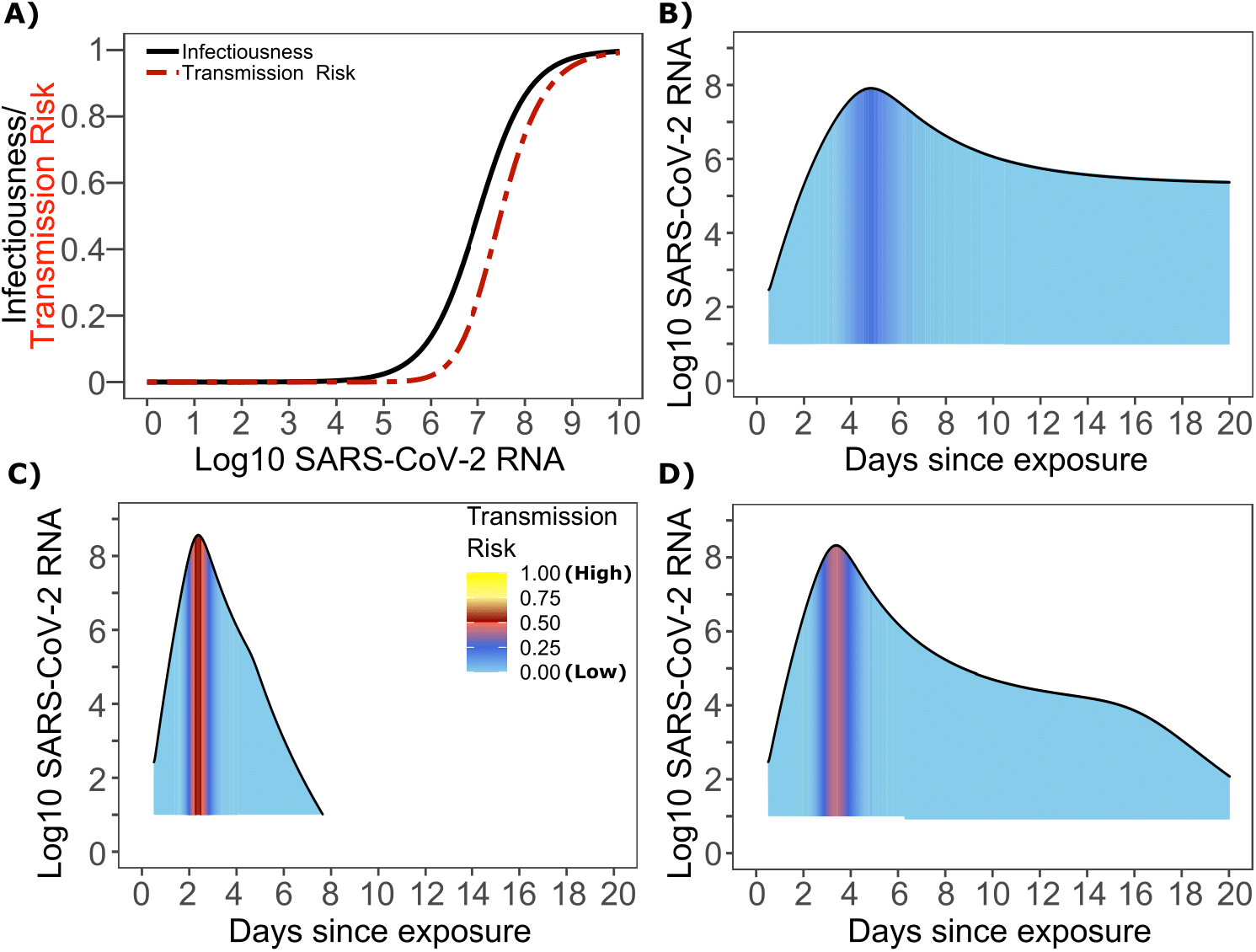
SARS-CoV-2 transmission probability as a function of shedding. **A**. Optimal infectious dose (ID) response curve (infection risk = *P_t_)* and transmission dose (TD) response curve (transmission risk = *P_t_ * P_t_)* curves for SARS-CoV-2. Transmission probability is a product of two probabilities, contagiousness and infectiousness (**Fig 1**). **B-D**. Three simulated viral shedding curves. Heat maps represent risk of transmission at each shedding timepoint given an exposed contact with an uninfected person at that time.

### Narrow duration of high infectivity during SARS-CoV-2 infection

We next plotted the probability of infection given an exposure to a transmitter. Under multiple shedding scenarios, the window of high probability transmission is limited to time points around peak viral load, and some heterogeneity in regard to peak infectivity is noted between people (**Fig 3b-d**). In general, infected persons are likely to be most infectious (i.e., above TD50) for a ~0.5-1.0-day period between days 2 and 6 after infection. We therefore conclude that the observed wide variance in serial interval (Fig 2c) results primarily from the possibility of highly discrepant incubation periods between the transmitter and infected person, rather than wide variability in shedding patterns across transmitters.

### Requirements for SARS CoV-2 super-spreader events

The solved value for exposed contact network heterogeneity (p) is 40 indicating high variability in day-to-day exposure contact rates (**Fig S3d**) with a high average number of exposed contacts per day (θ=4). We generated a heat map from our TD curve to identify conditions required for super-spreader events which included viral load exceeding 10^7^ SARS CoV-2 RNA copies and a high number of exposure contacts on that day. We observed an inflection point between 10^6^ and 10^7^ SARS CoV-2 RNA copies where large increases in the number of daily exposure contacts had a more limited impact on increasing the number of transmissions from a single person (**Fig 4a**). The exposure contact network occasionally resulted in days with ≥150 exposure contacts per day, which may allow an extremely high number of secondary infections from a single person (**Fig 4a**).

**Fig 4.**
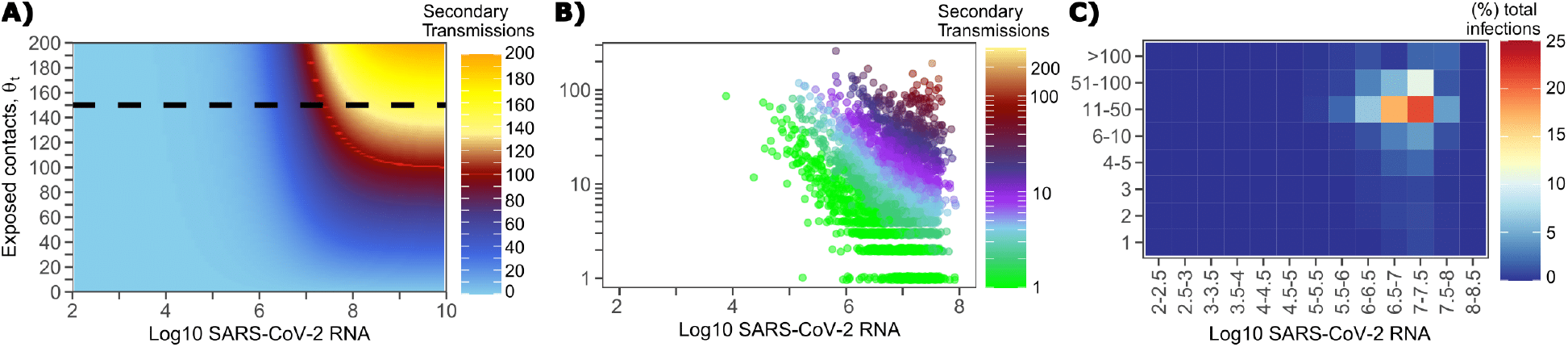
Conditional requirements for SARS-CoV-2 superspreading events. **A**. Heatmap demonstrating the maximum number of feasible secondary infections per day from a transmitter given an exposure viral load on log10 scale (x-axis) and number of exposed contacts per day (y-axis). The exposed contact network allows a maximum of 150 exposed contacts per day (black dotted line) which is sufficient for multiple transmissions from a single person per day. **B**. 10,000 simulated transmitters followed for 30 days. The white space is a parameter space with no transmissions. Each dot represents the number of secondary transmissions from a transmitter per day. Input variables are log10 SARS-CoV-2 on the start of that day and number of contact exposures per day for the transmitter. There are 1,154,001 total exposure contacts and 15,992 total infections. **C**. 10,000 simulated infections with percent of infections due to exposure viral load binned in intervals of 0.5 intervals on log10 scale (x-axis) and number of exposed contacts (y-axis).

We next plotted transmission events simulated on a daily basis over 30 days since infection, from 10,000 transmitters, according to viral load at exposure and number of exposure contacts on that day (Fig 4b). Secondary transmissions to only 1-3 people occurred almost exclusively with daily numbers of exposure contacts below 10 with any exposure viral load exceeding 10^6^ RNA copies or with higher numbers of exposure contacts per day and viral loads exceeding 10^5^ RNA copies. Massive super-spreader events with over 50 infected people almost always occurred at viral loads exceeding 10^7^ RNA copies with high levels of concurrent exposure contacts (**Fig 4b**).

We next identified that over 50% of secondary infections were associated with a transmitter who has a high number of exposed contacts (11-100 per day) and a viral load exceeding 10^6^ RNA copies (**Fig 4c**), which is the mechanistic underpinning of why ~70% of all secondary infections arose from transmitters who produced more than 10 secondary infections (**Table 1**).

### Model predicted individual R0 and serial intervals for influenza infection

A single model parameter set most closely reproduced empirically observed histograms and cumulative distribution functions for individual R0 and serial intervals for influenza: (*α, λ*, *τ*, *θ, ρ*) = (0.7, 10^5.5^, 0-0.5, 4, 1). ID50 values for influenza were lower than SARS CoV-2, but a direct comparison cannot be made because tissue culture infectious dose (TCID) has been more commonly used for measurements of influenza viral load, whereas viral RNA is used for SARS-CoV-2. Nevertheless, TCID is a closer measure of infectious virus and it is thus reasonable that ID50 based on TCID for influenza would be ~30-fold lower than ID50 based on total viral RNA (infectious and non-infectious virus) for SARS-CoV-2.^37^

The other notable difference was a considerably lower *ρ* value for influenza (**Fig S3b**), denoting much less heterogeneity in the number of exposure contacts per person while the average daily exposure contact was the same for both viruses (4 per day). The model captures the fact that 40% of influenza infected people do not transmit to anyone else and that each increase in the number of individual transmissions is associated with a lower probability (**Fig. 5a**). Relative to SARS-CoV-2, super-spreader events involving 5 or more people were predicted to be 5-fold less common overall and 10-fold less common among transmitters (~2% of all infected people and ~3% of transmitters) (**Fig. 5b, Table 1**). Super-spreaders defined as those infecting ≥5 individuals contributed to only ~10% to all transmissions (**Table 1**).

**Fig 5.**
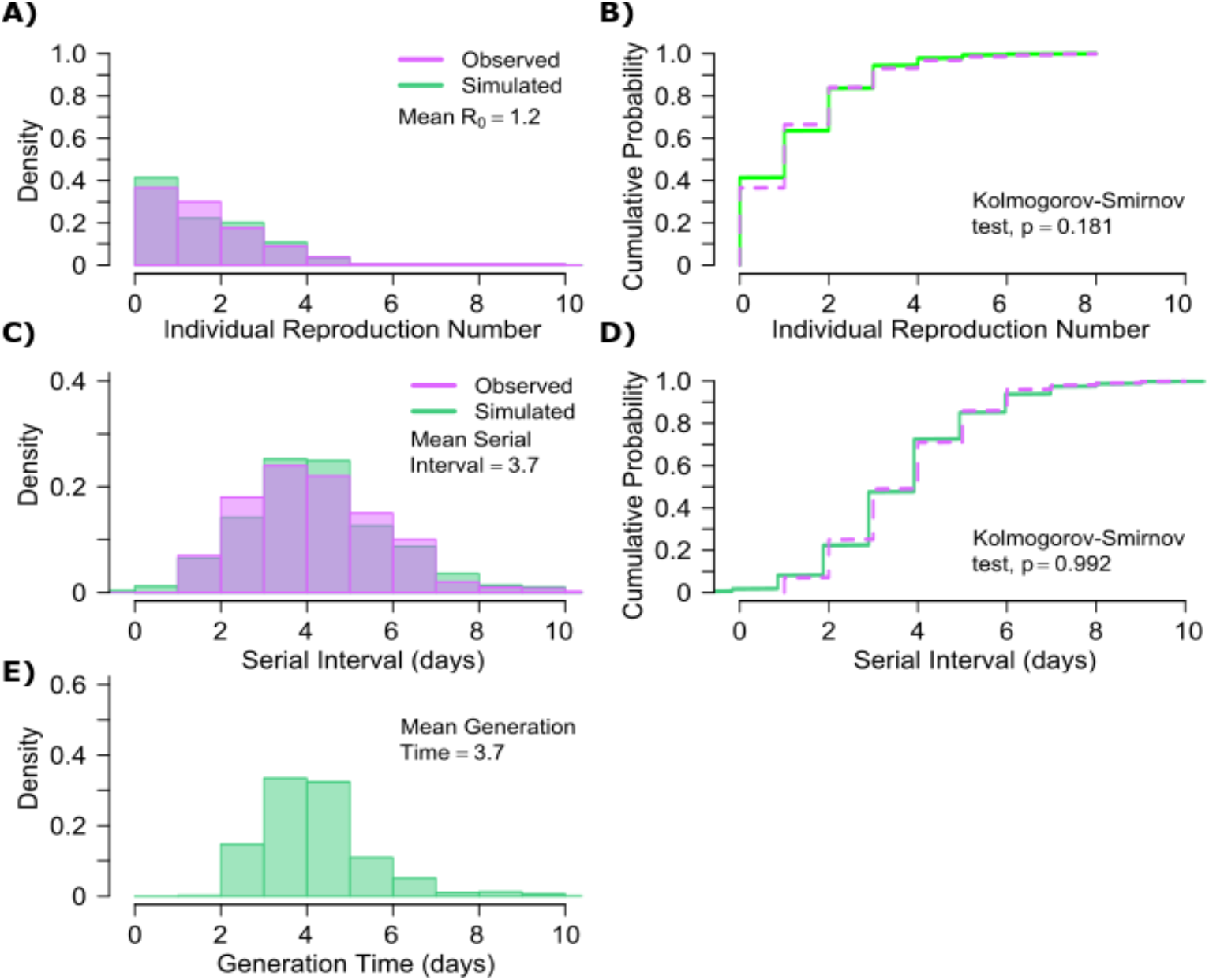
Influenza transmission model fit. **A.** Simulated and actual frequency histograms of individual R0 values, **B.** Simulated and actual cumulative distribution of individual R0 values. **C.** Simulated and actual frequency histograms of individual serial intervals, **D.** Simulated and actual cumulative distribution of individual serial intervals. **E.** Frequency distribution of simulated generation times.

The model also recapitulated the lower variance of serial interval for influenza relative to SARS-CoV-2 (**Fig 5c, d**). We next identified that the mean and variance of the serial interval provide good approximations of the mean and variance for generation time. A majority of generation times fell between 2 and 6 days (**Fig 5e**).

### Viral load thresholds for influenza transmission

Based on the optimized TD curve for influenza (**Fig 6a**), we next plotted the probability of infection given an exposure to an infected person. The TD50 for influenza was 10^6.1^ TCID/mL. Under various shedding scenarios, the window of high probability transmission was limited to time points around peak viral load (**Fig 6b-d**). In general, infected persons were likely to be most infectious (i.e., above TD50) for *α* ~0.5-1.0 days period. The observed low variance in serial interval (**Fig 5c**) resulted primarily from the narrow range of incubation periods within the transmitter and secondarily infected person, as well as the limited variability in shedding patterns across transmitters.

**Fig 6.**
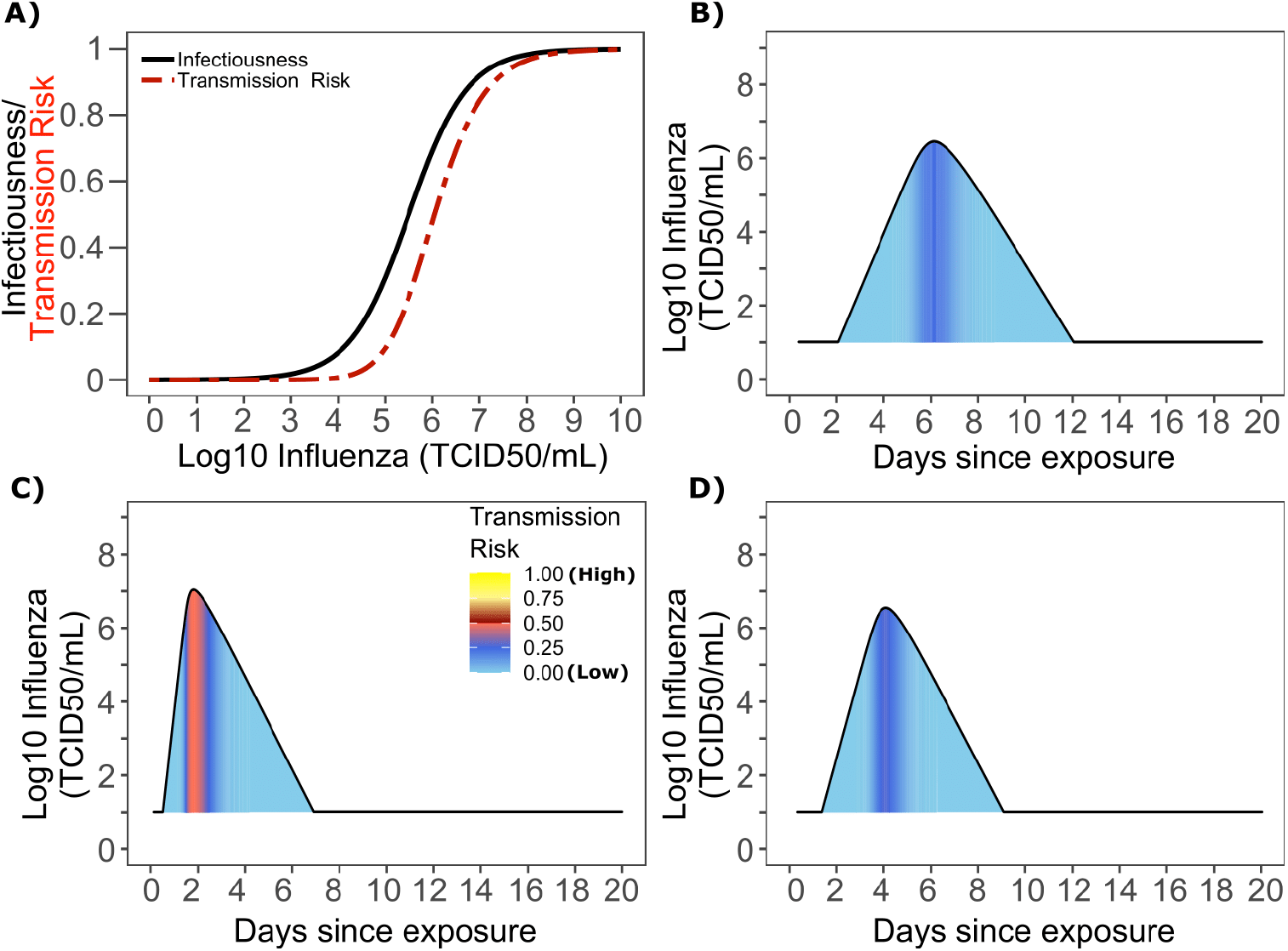
Influenza transmission probability as a function of shedding. **A.** Optimal infectious dose (ID) response curve (infection risk = *Pt)* and transmission dose (TD) response curve (transmission risk = *P_t_ * P_t_)* curves for influenza. Transmission probability is a product of two probabilities, contagiousness and infectiousness (Fig 1). **B-D**. Three simulated viral shedding curves. Heat maps represent risk of transmission at each shedding timepoint given an exposed contact with an uninfected person at that time.

### Determinants of influenza individual R0

We generated a heat map from our TD curve to identify conditions governing influenza transmission to multiple people including viral load exceeding 10^6^ influenza TCID and a high number of exposure contacts per day. The contact network never resulted in days with more than 15 exposure contacts per day, which severely limited the possible number of transmissions from a single person relative to SARS-CoV-2 (**Fig 7a, S3b**).

**Fig 7.**
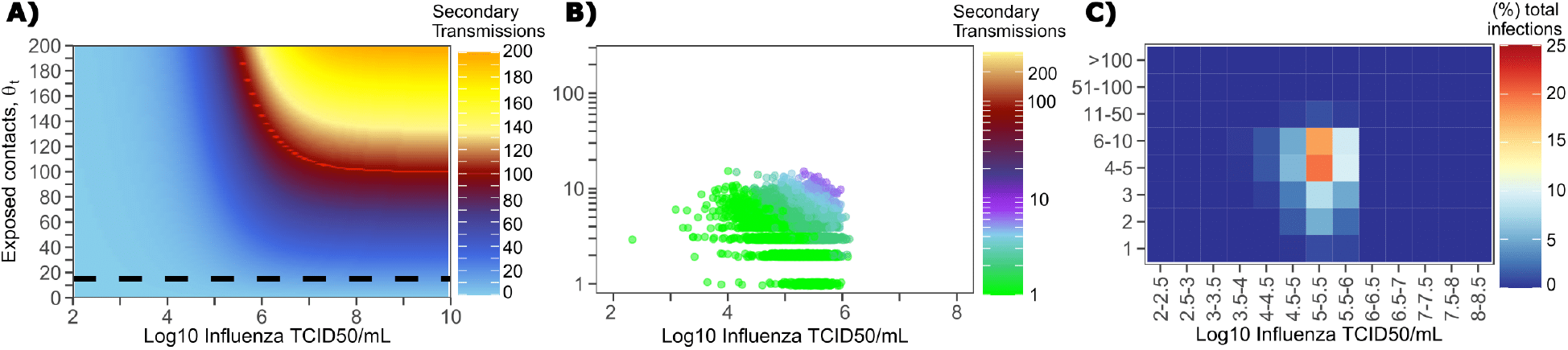
Conditional requirements for influenza super spreading events. **A**. Heatmap demonstrating the maximum number of secondary infections per day feasible from a transmitter given an exposure viral load on log10 scale (x-axis) and number of exposed contacts per day (y-axis). The exposed contact network allows a maximum of 15 exposed contacts per day (black dotted line) which is not sufficient for more than 15 transmissions from a single person per day. **B.** 10,000 simulated transmitters followed for 30 days. The white space is a parameter space with no transmissions. Each dot represents the number of secondary transmissions from a transmitter per day. Input variables are log10 influenza TCID on the start of that day and number of contact exposures per day for the transmitter. There are 1,239,984 total exposure contacts and 11,141 total infections. **C.** 10,000 simulated infections with percent of infections due to exposure viral load binned in intervals of 0.5 intervals on log10 scale (x-axis) and number of exposed contacts (y-axis).

We plotted transmission events simulated on a daily basis over 30 days since infection from 10,000 transmitters according to viral load at exposure and number of exposure contacts on that day (**Fig 7b**). Secondary transmissions to fewer than 5 people accounted for 90% of infections (**Table 1**) and occurred with fewer than 10 daily exposure contacts and exposure viral loads exceeding 10^4^ TCID. Small scale super-spreader events with 5-10 infected people almost always occurred at viral loads exceeding 10^5^ TCID with 5-10 concurrent exposure contacts (**Fig 7b**).

We next identified that over 50% of infections were associated with a transmitter who had fewer than 10 exposure contacts per day and a viral load exceeding 10^4.5^ TCID (**Fig 7c**), which is why no infected person ever transmitted to more than 10 other people (**Table 1**).

### Differing exposed contact distributions, rather than viral kinetics, explain SARS CoV-2 super-spreader events

We sought to explain why SARS-CoV-2 has a more over-dispersed distribution of individual R0 relative to influenza. To assess viral kinetics as a potential factor, we comparatively plotted transmission risk per exposure contact as a function of time since infection in 10,000 transmitters for each virus. The median per contact transmission risk was slightly higher for influenza; however, 75% and 95% transmission risks were marginally higher for SARS-CoV-2 compared to influenza with slightly higher peak transmission risk, and a longer tail of low transmission risk beyond 7 days (**Fig 8a**). The transmission risk was considerably higher for the 25% of simulated SARS-CoV-2 infections with the highest viral loads, suggesting that a substantial subset of infected people may be more pre-disposed to super-spreading. When plotted as time since onset of symptoms, the variability in transmission potential was considerably larger for persons with high SARS-CoV-2 viral load, owing to the variable incubation period of this virus (**Fig 8b**).

**Fig 8.**
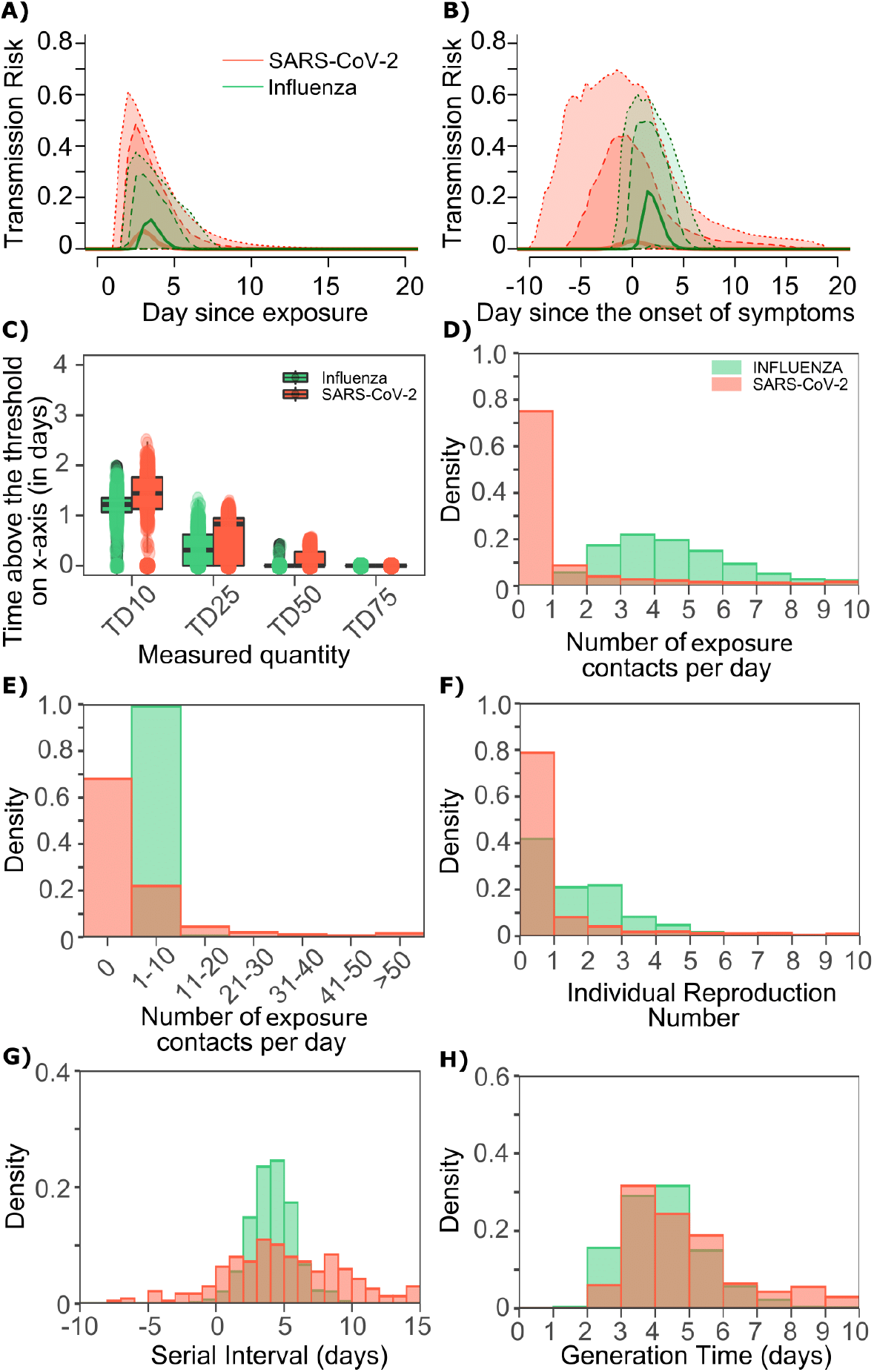
Differing transmission contact distributions, rather than viral kinetics explain SARS CoV-2 super spreader events. **A.** Simulated transmission risk dynamics for 10,000 infected persons with SARS-CoV-2 and influenza. Solid line is median transmission risk. Dark, dotted line is transmission risk of 75^th^ percentile viral loads, and light dotted line is transmission risk of 95^th^ percentile viral loads. **B.** Same as A but plotted as transmission risk since onset of symptoms. Highest transmission risk for SARS-Co-V-2 is pre-symptoms and for influenza is post symptoms. **C.** Boxplots of duration of time spent above TD10, TD25, TD50, TD75 and TD90 for 10,000 simulated SARS-CoV-2 and influenza shedding episodes. TD10, TD25, TD50, TD75 and TD90 are viral loads at which transmission probability is 10%, 25%, 50%, 75% and 90% respectively. The midlines are median values, boxes are interquartile ranges (IQR), and datapoints are outliers. Superimposed probability distributions of: **D & E.** number of transmission contacts per day, **F.** individual R0, **G.** serial interval and **H.** generation time for influenza and SARS-CoV-2.

The median duration of shedding over infectivity thresholds was short and nearly equivalent for both viruses. For SARS-CoV-2 and influenza, median [range] time above ID10 was 2.7 [0, 7] and 2.4 [1.6, 3.7] days respectively; median time above ID25 was 1.7 [0, 3] and 1.5 [0, 2.2] days respectively; median time above ID50 was 0.8 [0, 1.3] and 0 [0, 1.3] days respectively; median time above ID75 was 0 [0, 0.4] and 0 [0, 0] days respectively; median time above ID90 was 0 [0, 0] and 0 [0, 0] days respectively. ID10, ID25 and ID50 values were more variable across SARS-CoV-2 simulations due to a minority of trajectories with prolonged moderate viral loads.

For SARS-CoV-2 and influenza, median [range] time above TD10 was 1.4 [0, 2.5] and 1.2 [0, 2.0] days respectively; median time above TD25 was 0.8 [0, 1.3] and 0.3 [0, 1.3] days respectively; median time above TD50 was 0 [0, 0.5] and 0 [0, 0.4] days respectively; median time above TD75 was 0 [0, 0] and 0 [0, 0] days respectively. TD10, TD25 and TD50 values were more variable across SARS-CoV-2 simulations due to a minority of trajectories with prolonged moderate viral loads (**Fig 8c**).

We next plotted the frequency of exposure contacts per day for both viruses and noted a higher frequency of days with no exposed contacts (**Fig 8d**), but also a higher frequency of days with more than 10 exposure contacts (**Fig 8e**) for SARS-CoV-2 relative to influenza, despite an equivalent mean number of daily exposure contacts. To confirm that this distribution drives the different observed distributions of individual R0 values (**Fig 8f**), we simulated SARS-CoV-2 infection with an assumed *ρ*=1 and generated a distribution of individual R0 similar to that of influenza (Fig S6a). Similarly, we simulated influenza infection with an assumed *ρ*=40 and generated a distribution of individual R0 similar to that of SARS-CoV-2 (**Fig S6b**). Under all scenarios, predicted distributions of serial interval (**Fig 8g, Fig S6**) and generation time (**Fig 8h, Fig S6**) were unchanged by shifts in the exposed contact network.

### Projections of targeted physical distancing

Physical distancing is a strategy to decrease R0. We simulated a decrease in the contact rate uniformly across the population and noted a decrease in population R0 (**Fig S7a**) as well the percent of infected people who will transmit (**Fig 7b**) and become super-spreaders (**Fig S7c-d**). An approximately 40% decrease in the average exposed contact rate decreased R0 below 1 (**Fig S6a**). We further investigated whether lowering contact rate among larger groups only, in particular by banning exposure events with a high number of exposure contacts, could control the epidemic. We identified that limiting exposure contacts to no more than 5 per day is nearly equivalent to limiting exposure contacts altogether and that only a small decrease in mean exposure contact rate can achieve R0<1 if exposure events with less than 20 contacts are eliminated (**Fig S8**).

### Pre-symptomatic transmission and super-spreading risk

Much of the highest transmission risk for SARS-CoV-2 exists in the pre-symptomatic phase (**Fig 8b**) which explains why 62% of simulated transmissions occurred in the pre-symptomatic phase for SARS-CoV-2, compared to 10% for influenza. Similarly, 62% and 21% of SARS-CoV-2 and influenza super-spreader events with secondary transmissions ≥5 and 39% of SARS-CoV-2 super-spreader events with secondary transmissions R0≥10 fell in the pre-symptomatic period.

## Discussion

Our results demonstrate that SARS-CoV-2 shedding kinetics are directly linked to the virus’ most fundamental epidemiologic properties. First, we identify a transmission dose response curve which specifies that a nasal viral load below 10^5^ RNA copies is unlikely to commonly result in transmission. For SARS-CoV-2, this threshold is consistent with the overall rarity of positive cultures at these levels.^37^ We also predict a relatively steep TD curve such that transmission becomes much more likely when shedding exceeds 10^8^ viral RNA copies and there is an exposure contact between an infected person and susceptible person. The amount of viral RNA can be roughly converted to the probability of a positive viral culture which approximates infectiousness. Our results therefore have relevance for dosing of SARS-CoV-2 in human challenge experiments that are being considered for vaccine trials.

While the duration of shedding for SARS-CoV-2 is often three weeks or longer,^11,12^ we predict that the duration of shedding above thresholds required for a moderate probability of transmission per contact is much shorter, often less than half a day, and is comparable to that of influenza. While transmission after the first week of infection is quite rare, our model is consistent with the observation that transmissions commonly occur during the pre-symptomatic phase of infection,^2^ given the highly variable incubation period associated with SARS-CoV-2.

The observed high heterogeneity in serial interval is attributable almost entirely to the variable nature of the incubation period, rather than transmission occurring extremely late after infection. While our estimate for mean generation time is equivalent to that of mean serial interval, it is notable that the range of SARS-CoV-2 serial intervals is much wider than the range of generation times. This result is evident even though we built substantial heterogeneity into our viral shedding curves beyond that observed in the somewhat limited existing shedding data.

The finding of limited duration of SARS-CoV-2 infectivity has practical implications. First, considerable resources are being used in hospitals and skilled nursing facilities to isolate patients with persistent SARS-CoV-2 shedding. We propose that a low nasal viral load, particularly during late infection, need not justify full patient isolation procedures in the absence of aerosolizing procedures. This observation could save substantial hospital resources and valuable isolation beds during subsequent waves of infection. Similar considerations are relevant for employees wishing to return to work. Our results also suggest that time since first positive test may be predictive of lack of contagion, though more viral load kinetic studies will be needed to confirm the existing observation that viral loads after a week of infection are usually low and associated with negative viral cultures.^37^ Finally, our conclusions are supportive of rapid, less sensitive assays which are more likely to detect infection at periods of contagion.^43^

Many of these conclusions, including specific viral load thresholds for transmission, a steep dose response curve and a maximum 2-day duration of contagion within an infected individual are equally relevant for influenza infection. One important difference is that incubation periods for influenza are far less variable which means that at the individual level, the serial interval is much more likely to be predictive of the generation time.

Another finding is that SARS-CoV-2 super-spreading events are dependent on a large number of exposure contacts during the relatively narrow 1-2 days window during which a ~25% subset of infected people is shedding at extremely high levels above the TD50. Because we predict that super-spreader potential may be somewhat of a generalized property of infection, rather than a characteristic of a tiny subset of infected people, this result also has practical implications. A common experience during the pandemic has been early identification of a cluster of infected people within a specific confined environment such as a senior living home, crowded work environment, athletic team, or restaurant. Our results demonstrate that newly diagnosed people within small clusters may be past the peak of their super-spreading potential. At this stage, many more infections have often been established and drastic quarantine procedures should be considered. Other undiagnosed, pre-symptomatic infected people may have super-spreader potential while the known infected person is no longer contagious, highlighting the importance of effective contact tracing.

At the prevention level, school opening and work opening strategies should focus on severely limiting the possible number of exposure contacts per day. Where large numbers of exposure contacts are unavoidable, mandatory masking policies, perhaps with N95 masks that may more significantly lower exposure viral loads should be considered.^23^

Influenza infection is much less predisposed to super-spreader events than SARS-CoV-2. Yet, influenza shedding at levels above those required for a high probability of transmission occurs with only slightly lower frequency. Therefore, the markedly different probability of super-spreader events between the two viruses is unlikely to relate to different viral host kinetics, despite the fact that the overall duration of SARS-CoV-2 shedding exceeds duration of influenza shedding often by more than two weeks.

Rather, our analysis suggests that the exposure contact networks of SARS-CoV-2 transmitters are highly variable relative to those of influenza. One possible explanation underlying this finding is that SARS-CoV-2 is more predisposed to airborne transmission than influenza.^44^ Here our precise definition of an exposure contact (sufficient contact between a transmitter and an uninfected person to potentially allow transmission) is of high relevance. Our result suggests that a SARS-CoV-2 infected person in a crowded, poorly ventilated room, may generate more exposure contacts than an influenza infected person in the same room, likely based on wider dispersal and / or longer airborne survival of the virus. Thus, our results suggest a possible downstream quantitative effect of airborne transmission on SARS-CoV-2 epidemiology. Another possibly important variable is that pre-symptomatic transmission, which is a common feature of SARS-CoV-2 may predispose to multiple transmissions. This prediction reinforces current public health recommendation to avoid crowded indoor spaces with poor air recirculation.

On the other hand, a much higher proportion of SARS-CoV-2 infected people than influenza infected people do not transmit at all. This result lacks a clear mechanistic explanation but may imply that aerosolization occurs only in a subset of infected people. One theoretical explanation is that high viral load shedding in the pre-symptomatic phase is defined by lack of cough or sneeze leading to limited spatial diffusion of virus. Alternatively, it is also possible that a proportion of infected people never shed virus at high enough viral loads to allow efficient transmission. This possibility speaks to the need for more quantitative viral load data gathered during the initial stages of infection.

Age cohort structure differs between the two infections, with a lower proportion of observed pediatric infections for SARS-CoV-2. If adults have more high exposure events than children, then this could also explain super-spreader events. We are less enthusiastic about this hypothesis. First, SARS-CoV-2 super-spreader events have occurred in schools and camps and would likely be more common in the absence of widespread global school closures in high prevalence regions. Second, a sufficient proportion of influenza cases occur in adults to rule out the presence of frequent large super-spreading events in this population.

Our analysis has important limitations. First, exposure contacts were assumed to be homogeneous and we do not capture the volume of the exposing aerosol or droplet. For instance, if a large-volume droplet contains ten times more viral particles than an aerosol droplet, then the exposure could be dictated by this volume as well as the viral load of the potential transmitter. It is possible that under rare circumstances with extremely high-volume exposures, even persons with extremely low viral loads may transmit. Second, based on the quality of available data, we fit our models for SARS-CoV-2 and influenza to viral RNA and viral culture respectively. Existing data suggest that kinetics of viral RNA and culture are similar during both infections, with culture having lower sensitivity to detect virus.^37^ Third, our intra-host model of SARS-CoV-2 was fit to heterogeneous data with different sampling techniques and PCR assays.^24^ Moreover, R0 estimates have varied across the globe. Our estimates of TD50 are necessarily imprecise based on available data and should serve only as a conservative benchmark. Most importantly, we cannot rule out the possibility that a small minority of infected people shed at sufficient levels for transmission for much longer than has been observed to date. Fourth, contagiousness could have different dose response dynamics than viral load dependent infectiousness and may require investigation in the future upon the availability of epidemiologically relevant additional data. Finally, the model is intended to capture a general property of SARS-CoV-2 infection but is not specific for local epidemics. The degree of R0 overdispersion in various countries and regions is likely to vary dramatically according to numerous factors related to social contact networks that are not explicitly captured in our model.

In conclusion, fundamental epidemiologic features of SARS-CoV-2 and influenza infections can be directly related to viral shedding patterns in the upper airway as well as the nature of exposure contact networks. We contend that this information should be leveraged for more nuanced public health practice in the next phase of the pandemic.

## Methods

### SARS-CoV-2-within-host model

To simulate SARS-CoV-2 shedding dynamics, we employed our previously-described viral infection model.^24^ In this model, susceptible cells (S) after coming into contact with SARS-CoV-2 (V) become infected at rate *βVS*. The infected cells *(I)* produce new virus at a per-capita rate *π*. The model also includes the clearance of infected cells in two ways: (1) by an innate response with density dependent rate *δI^k^*; and (2) an acquired response with rate 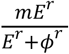 mediated by SARS-CoV-2-specific effector cells *(E)*. The clearance mediated by innate immunity depends on the infected cell density and is controlled by the exponent *k*. The Hill coefficient *r* parameterizes the nonlinearity of the second response and allows for rapid saturation of the killing. Parameter *ϕ* defines the effector cell level by which killing of infected cells by *E* is half maximal.

In the model, SARS-CoV-2-specific effector cells rise after 2 stages from precursors cells *(M*_1_ and *M*_2_). The first precursor cell compartment *(M*_1_) proliferates in the presence of infection with rate *ωIM*_1_ and differentiates into the effector cell at a per capita rate *q* during the next intermediate stage. Finally, effector cells die at rate *δ_E_*. The model is expressed as a system of ordinary differential equations:

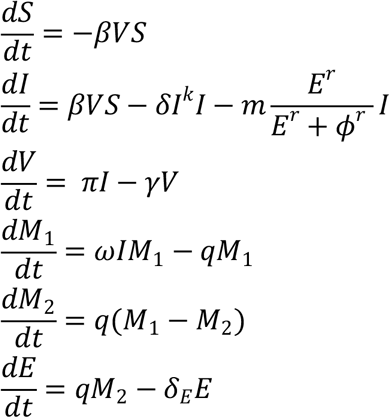

We assumed *S*(0) = 10^7^ cells/mL, *I*(0) = 1 cells/mL, 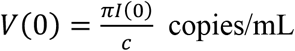, *M*_1_(0) = 1, M_2_(0) = 0 and *E*_0_ = 0.

When we introduce simulated heterogeneity in cases of SARS-CoV-2 (by increasing the standard deviation of the random effects of parameters *(β* by 20, *δ* by 2, *k* by 2 and *π* by 5 in the original distribution from^24^), some of the viral shedding curves suggest that viral shedding could continue for long period (over 6 weeks). Indeed, while median viral shedding duration has been estimated at 12-20 days, shedding for many months is also observed commonly.^45^ We assumed that viral loads after day 20 drop to a exposure-level viral load level (i.e., *V*(0)) as most viral shedding observed after this point is transient and at an extremely low viral load.^46^ The population distribution of parameters to simulate artificial SARS-CoV-2 viral shedding dynamics is provided in **Table S1**.

### Influenza within-host model

To simulate viral shedding dynamics of influenza viral, we employ a model^38^ that is a simplified version of the viral dynamics model presented for SARS-CoV-2. This model assumes *k* = 0 and *m* = 0 and can be expressed as a system of ordinary differential equations:

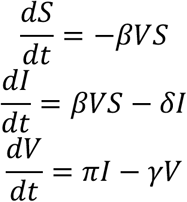

Following this model,^38^ we assumed *S*(0) = 4 × 10^4^ cells/mL, *I*(0) = 1 cells/mL, 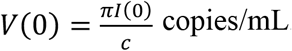. To simulate artificial influenza viral shedding dynamics, we assumed the population distribution of parameters *Log*_10_*(β), Log*_10_*(π), Log*_10_*(γ)* and *Log*_10_*(δ)* are −4.56 (0.17), −1.98 (0.14), 0.47 (0.03) and 0.60 (0.06), respectively.

### Dose-response model

For both viruses, to estimate the infectiousness *P_t_* [*V*(*t*)] based on viral loads *V(t)*, we employed the function, 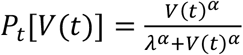. Here, *λ* is the infectivity parameter that represents the viral load that corresponds to 50% infectiousness and 50% contagiousness, and *α* is the Hill coefficient that controls the slope of the dose-response curve.

### Transmission Model and Reproduction number

Our transmission model assumes that only some contacts of an infected individual with viral load dependent infectiousness are physically exposed to the virus (defined as exposure contacts), that only some exposure contacts have virus passaged to their airways (contagiousness) and that only some exposed contacts with virus in their airways become secondarily infected (successful secondary infection). Contagiousness and infectiousness are then treated as viral load dependent multiplicative probabilities with transmission risk for a single exposure contact being the product. Contagiousness is considered to be viral load dependent based on the concept that a transmitter’s dispersal cloud of virus is more likely to prove contagious at higher viral load, which is entirely separate from viral infectivity within the airway once a virus contacts the surface of susceptible cells.

We next assume that the total exposed contacts within a time step (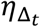) is gamma distributed, i.e. 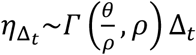, using the average daily contact rates (*θ*) and the dispersion parameter (*ρ*). To obtain the true number of exposure contacts with airway exposure to virus, we simply multiply the contagiousness of the transmitter with the total exposed contacts within a time step (i.e., 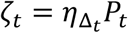).

Transmissions within a time step are simulated stochastically using time-dependent viral load to determine infectiousness (*P_t_*). Successful transmission is modelled stochastically by drawing a random uniform variable (*U*(0,1)) and comparing it with infectiousness of the transmitter. In the case of successful transmission, the number of secondary infections within that time step (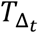) is obtained by the product of the infectiousness *(P_t_*) and the number of exposure contacts drawn from the gamma distribution *((ζ_t_*). In other words, the number of secondary infections for a time step is 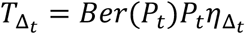. If we disregard contagiousness by assuming *P_t_* = 1 in *ζ_t_*, we identify that there are little to no differences on overall results other than the emergent TD curve and optimal parameter set describing dose-response curve and exposed contact network, which no longer agrees as closely with in vitro probability of positive virus culture (**Fig S5**).^37^

We obtain the number of secondary infections from a transmitter on a daily basis noting that viral load, and subsequent risk, does not change substantially within a day. We then summed up the number of secondary infections over 30 days since the time of exposure to obtain the individual reproduction number, i.e. 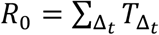.

### Serial interval and generation time

We further assume that upon successful infection, it takes *τ* days for the virus to move within-host, reach infection site and produce the first infected cell. To calculate serial interval (time between the onset of symptoms of transmitter and secondarily infected person), we sample the incubation period in the transmitter and in the secondarily infected person from a gamma distribution with a shape described in the **Fig S4**.^3,30^ In cases in which symptom onset in the newly infected person precedes symptom onset in the transmitter, the serial interval is negative; otherwise, serial interval is non-negative. We calculate generation time as the difference between the time of infection of transmitter and the time of infection of secondarily infected person.

### Individual R0 and serial interval data for model fitting

There is abundance of data confirming over-dispersed R0 for SARS-CoV-2. From contact tracing of 391 SARS-CoV-2 cases in Shenzhen, China, 1286 close contacts were identified: the distribution of individual R0 values in this cohort was highly over-dispersed, with 80% of secondary infections being caused by 8-9% of infected people.^6^ In another study, authors analyzed the contact/travel history of 135 infected cases in Tianjin, China and determined heterogeneity in the individual R0.^34^ Another contract tracing study also identified and characterized SARS-CoV-2 clusters in Hong Kong and estimated that 20% of cases were responsible for 80% of local transmission.^35^

A modeling study that simulated observed outbreak sizes in ~40 affected countries during the early phase of epidemics also confirmed that ~80% of secondary transmissions may have been caused by a small fraction of infectious individuals (~10%).^4^ The latter study provided the distribution of individual R0 (**Fig 2A**) that we employed for fitting purposes. Using the data on 468 COVID-19 transmission events reported in mainland China, Du et al. estimated the mean serial interval as well as the distribution of serial interval (**Fig 2C**).^31^ We employed this data for fitting purposes.

The cumulative distribution function of individual R0 for influenza was obtained from a modeling study that simulated the transmission dynamics of seasonal influenza in Switzerland from 2003 to 2015.^10^ We picked the parameters mean R0=1.26 and dispersion parameter=2.36 in the negative binomial distribution that corresponded to the 2008-2009 influenza A H1N1 pandemic.^10^ Another modeling study that simulated the age-specific cumulative incidence of 2009 H1N1 influenza in 8 Southern Hemisphere Countries yielded similar results.^40^ By following the household members of index cases, a study estimated the cumulative distribution of serial interval based on symptom-onset times from 14 transmission pairs.^9^ We employed these cumulative distribution functions of individual R0 and serial interval of influenza for fitting purposes.

### Fitting procedure

To estimate the values of unknown parameters in cases of SARS-CoV-2, we performed a grid search comprehensively exploring a total of ~500,000 combinations of 5 parameters taking the following values,

i. *τ* ∊ [0.5,1,2, 3] days,
ii. *α* ∊ [0.01, 0.1, 0.2,0.3,0.4, 0.5, 0.6, 0.7,0.8,0.9,1.0, 2.0, 3.0,4.0, 5.0,10.0]
iii. *X* ∊ [10^0^,10^0.5^,10^1.0^ …,10^8^]
iv. *θ* ∊ [0.1, 0.2, 0.3, 0.4, 0.5, 0.6, 0.7, 0.8, 0.9, 1.0, 2.0, 3.0, 4.0, 5.0, 10.0, 20.0, 50.0].
v. *ρ* ∊ [0.0001, 0.001, 0.01, 0.1, 0.2, 0.3, 0.4, 0.5, 0.6, 0.7, 0.8, 0.9, 1.0, 2.0, 5.0,10.0, 20.0, 30.0,40.0, 50.0, 75.0, 100, 200, 500].

The parameter sets of (*λ*, *τ*, *α*, *θ*) were simulated for 1000 infected individuals to determine how well each set generates the summary statistics of mean R0, mean SI and the R0 histograms by following a procedure explained in **Fig S1** and below:

#### Step A

1. Simulate viral load *V(t*) of 1,000 simulated infected individuals using **Eq. 1**
2. For each combination of (λ, *τ*, *α*, *θ*)
  a. For each time step Δ*_t_*
    i. Compute *P_t_ [V(t); λ*, *α]*
    ii. Draw 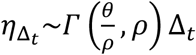
    iii. Calculate 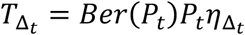
  b. Calculate 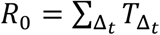
    i. Check if calculated mean *R*_0_ is in the range:^3,31^
  c. Calculate Serial Interval based on *τ* and incubation period
    i. Check if calculated *SI* is in the range in:^3,31,33^

#### Step B

1. If the parameter combination in Step A satisfy the criteria, then
  i. Compute RSS for the obtained *R*_0_ and histogram from:^4,6,34,36^ [Ref]

We visually checked whether our dose-response curve matched the observed probability of positive virus culture.^37^ We assumed that viral loads derived from positive culture^37^ can be considered equivalent to viral loads in the within-host model if divided by a positive integer. We identified an integer of 25 to provide closest fit to the empirical data (**Fig S5**).

We performed a global sensitivity analysis to identify which parameter variability accounted for fit to different components of the data. Only narrow ranges of *λ* permitted close fit to the mean of R0 and distribution functions of individual R0 (**Fig S9**), while a specific value for a was necessary to fit to mean serial interval and distribution functions of individual R0 (**Fig S9**). Only narrow ranges of θ permitted close fit to the mean of R0 and distribution functions of individual R0 (**Fig S10**), while a specific value for ρ was necessary to fit to distribution functions of individual R0 (**Fig S10**).

> To obtain TD50 *(λ_T_*) based on ID50 *(λ)*, we use the relation
>
> 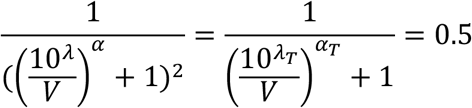
>
> From solving the second half 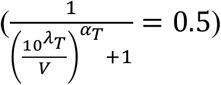, we get
>
> 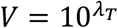
>
> Substituting *V* = 10*^λT^* in the first-half, we have
>
> 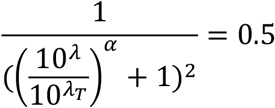
>
> Or, 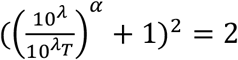
>
> Or, 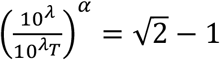
>
> Or, 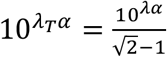
>
> Or, 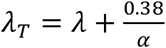

## Data Availability

All code used to generate our results will be made available on gut hub and can be accessed by emailing Dr. Schiffer

## Acknowledgements

We are grateful to study participants from around the globe who donated critical virologic data early during the pandemic. We thank Jeroen van Kampen and Marion Koopmans for helpful discussions.

## Author contributions

J.T.S. and B.M. conceived the study. A.G., E.F.C., B.M. and D.B.R. assembled data, wrote all code, performed all calculations and derivations, ran the models, and analyzed output data. J.T.S. wrote the manuscript with contributions from all other authors.

## Data and materials availability

The original data and code is shared at: https://github.com/ashish2goyal/SARS_CoV_2_Super_Spreader_Event

## Supplementary Materials

**Fig. S1.**
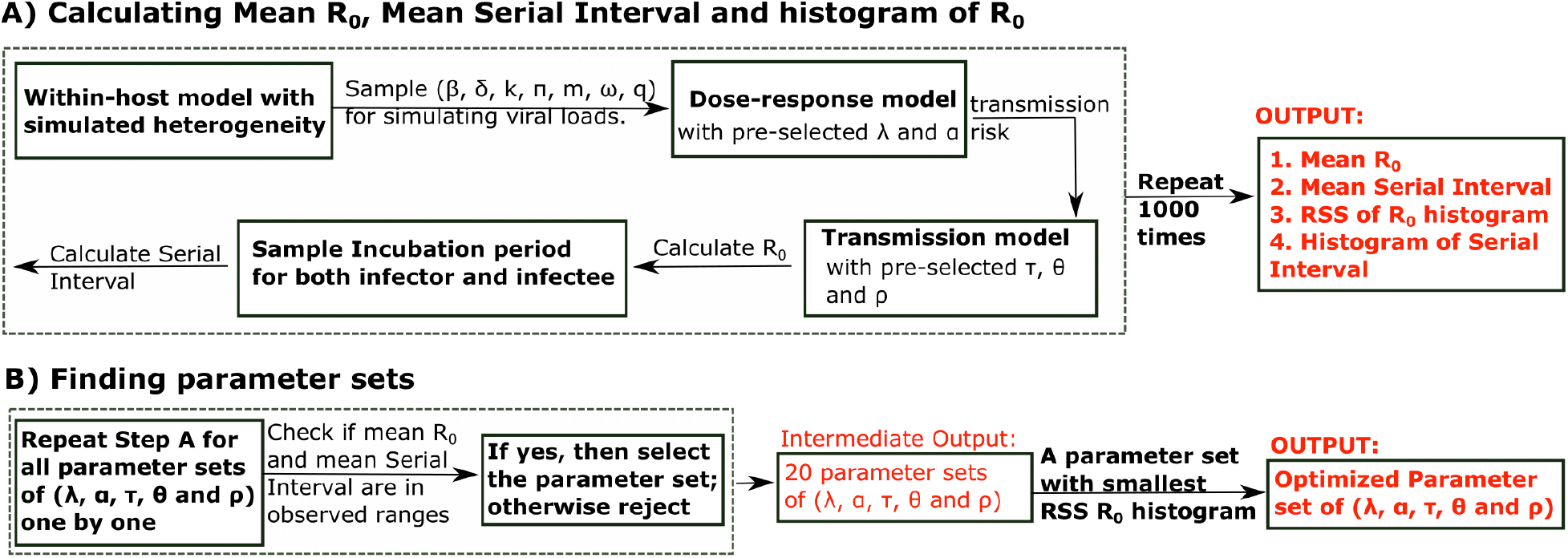
Mathematical model workflow.

**Fig. S2.**
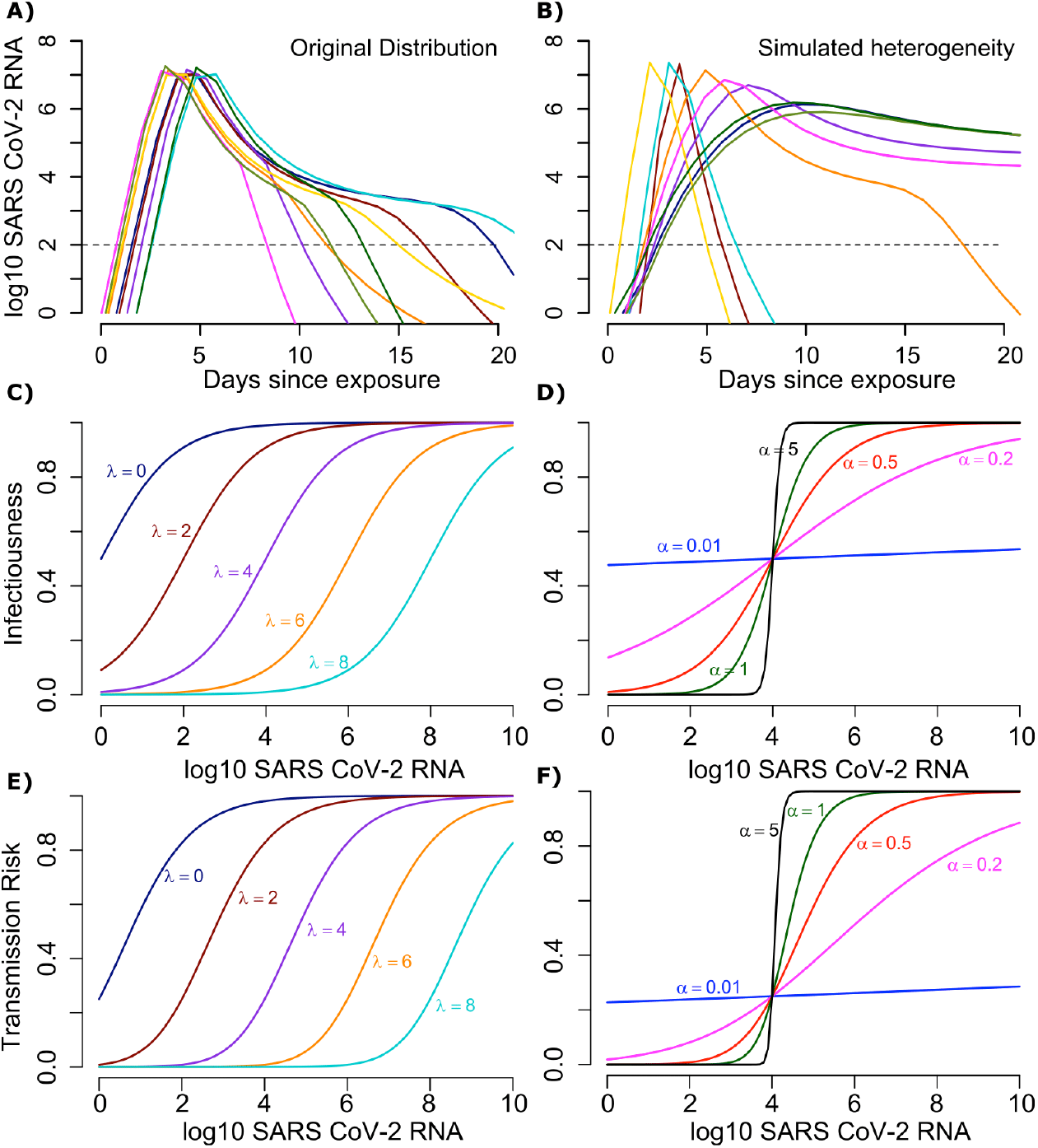
Mathematical model of SARS-CoV-2 transmission dynamics. **A**. Simulated viral load shedding tracings of possible transmitters. **B**. Simulated viral load shedding with imputed heterogeneity. **C**. Simulated infection dose (ID) response curves with variance in infectivity (ID50) and **D**. dose response slopes. **E**. Simulated transmission dose (TD) response curves with variance in infectivity (TD50) and **F**. dose response slopes. The TD response curve is a product of the infection and contagion dose response curves.

**Fig. S3.**
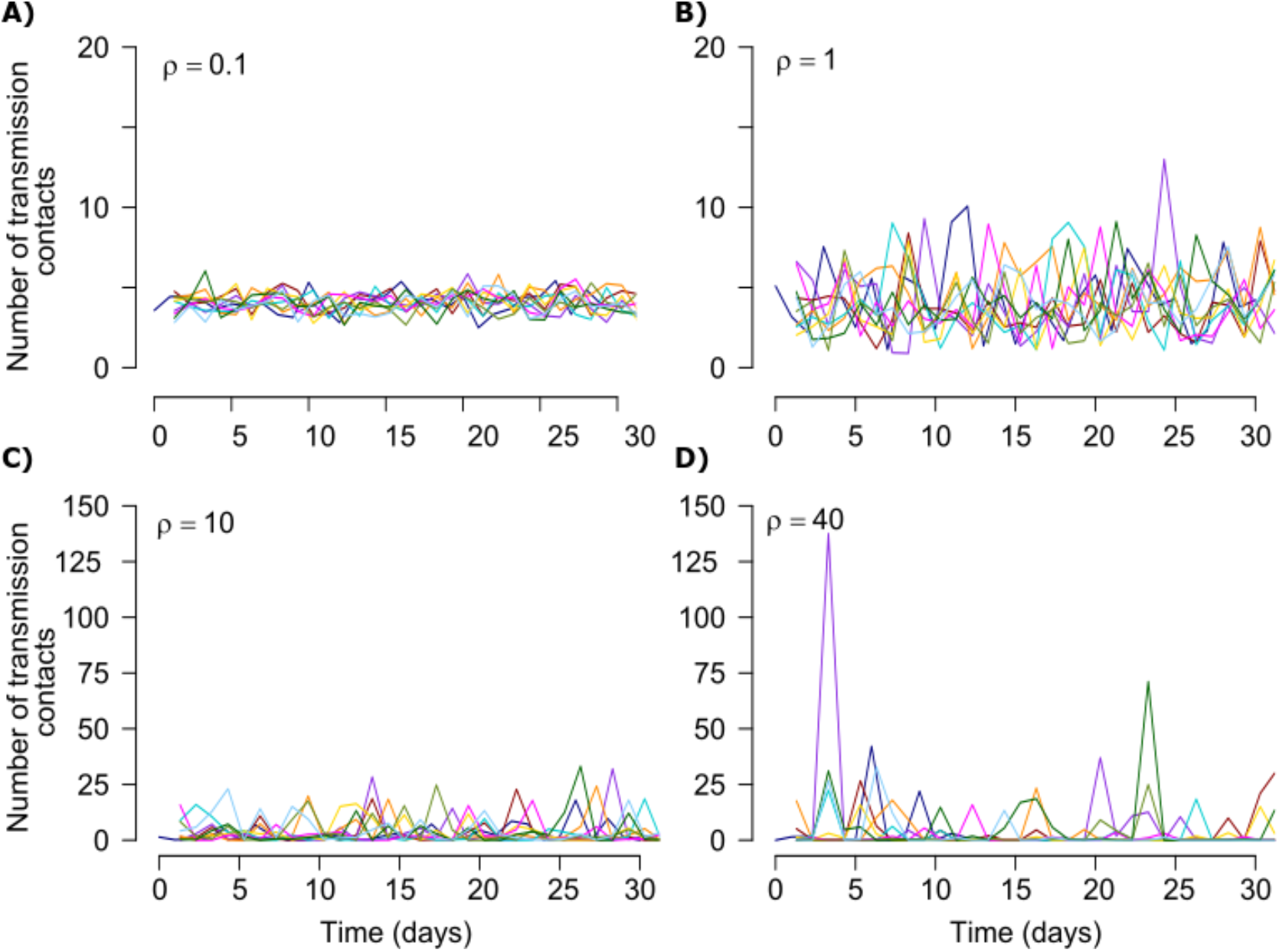
Stochastic simulations of exposed contact frequency for varying dispersion (ρ). The average number of exposed contacts is 4 per day in each example with imputed daily heterogeneity based on an elevated value of ρ from a gamma distribution~Γ(4/ρ, ρ).

**Fig. S4.**
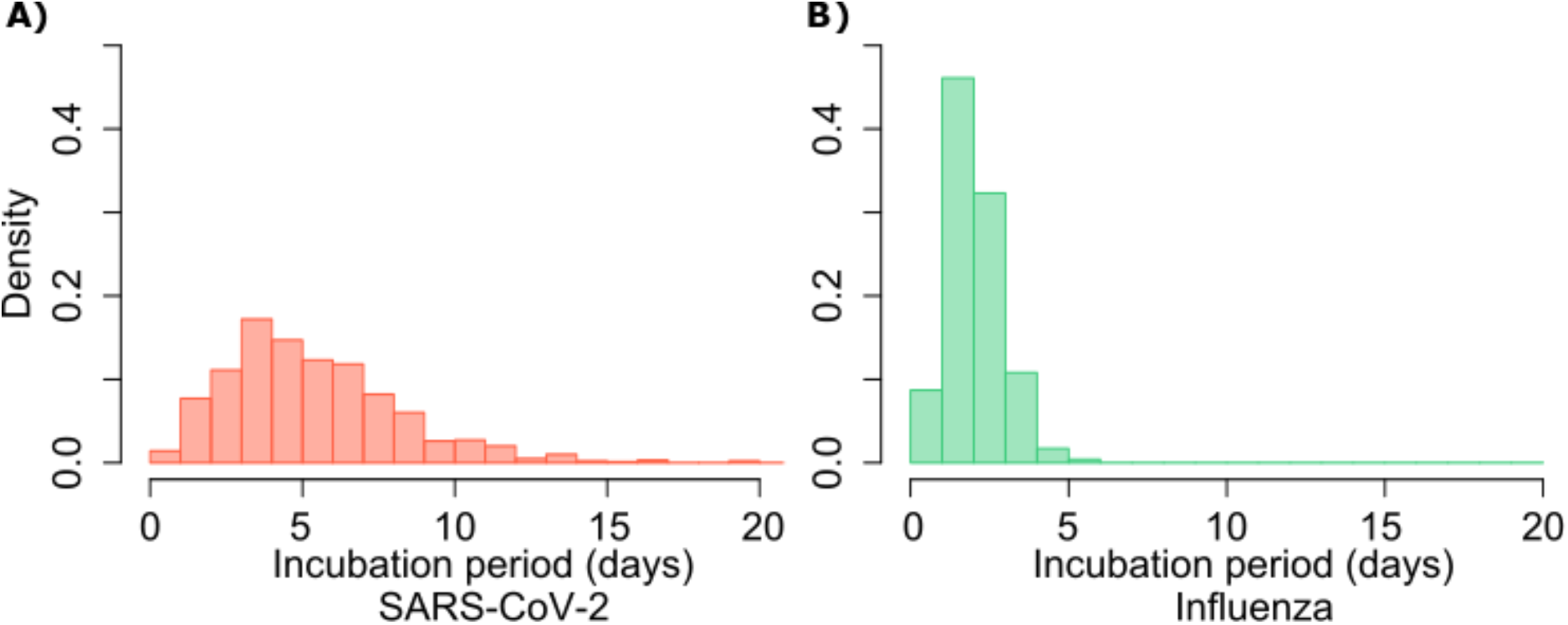
Gamma distribution functions of incubation periods. **A**. SARS-CoV-2 (mean 5.2 days, shape parameter =3.45 and rate =0.66) and **B**. influenza (mean 2 days, shape parameter=6.25 and scale parameter=0.32).

**Fig. S5.**
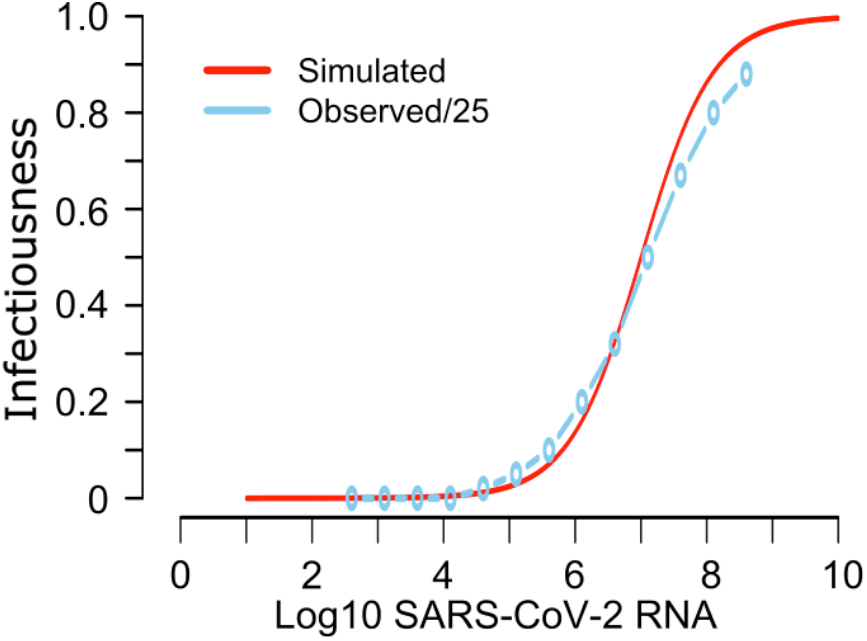
Mathematical model recapitulation of relationship between SARS-CoV-2 viral load and viral culture. In a clinical study, probability of positive viral culture was projected against SARS-CoV-2 RNA (https://www.medrxiv.org/content/10.1101/2020.06.08.20125310v1). When we divided these PCR values by 25 (light blue line), we identified high similarity between the clinical data and our projected infectiousness dose response curve (red line).

**Fig. S6.**
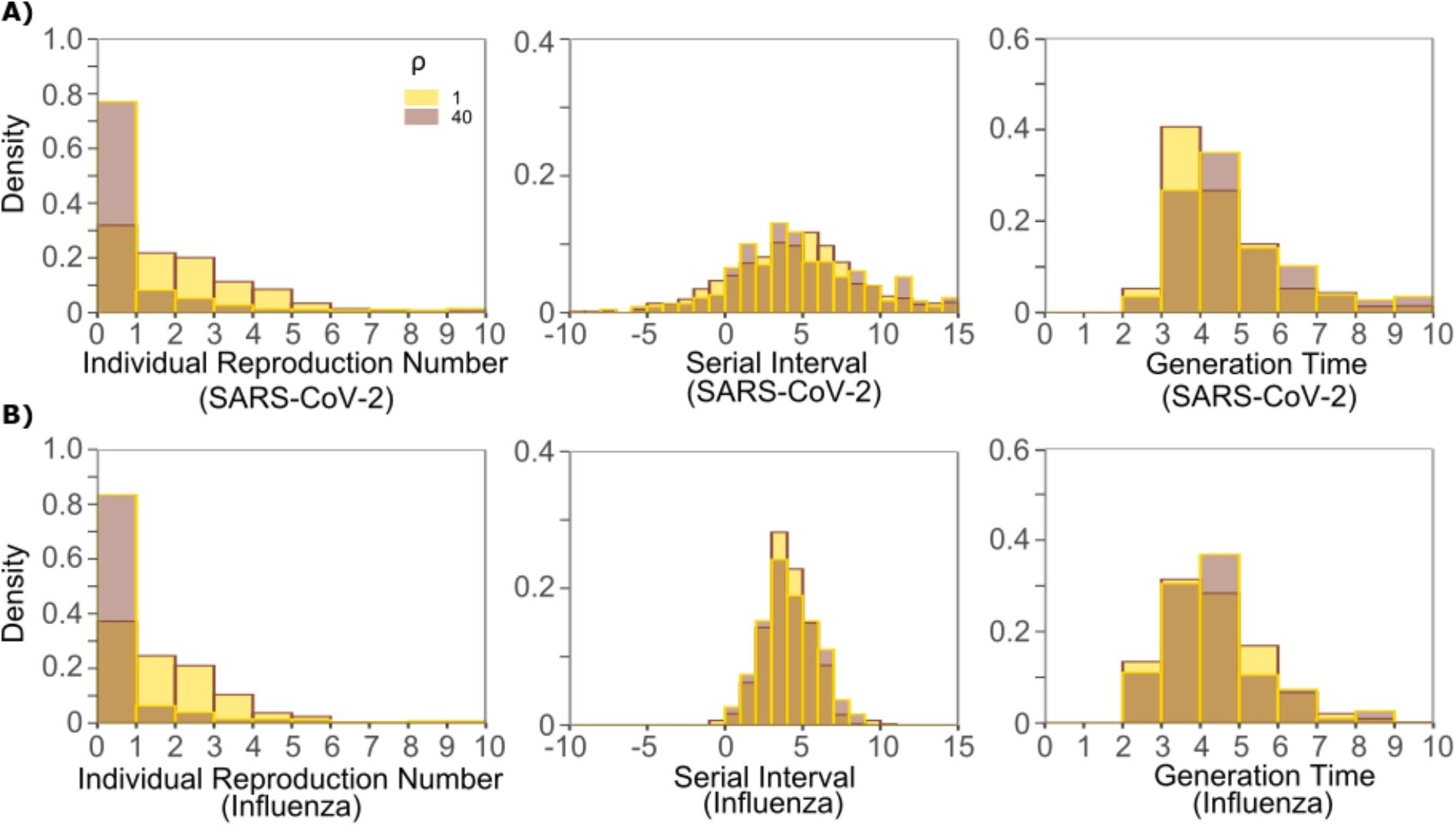
Impact of changes in contact network heterogeneity on individual R0, serial interval, and generation time. **A**. SARS-CoV-2, and **B**. influenza. Lowering exposed contact network heterogeneity to levels observed with influenza decreases SARS-CoV-2 individual R0 over-dispersion. Increasing exposed contact network heterogeneity to levels observed with SARS-CoV-2 increases influenza R0 over-dispersion. Neither change impacts observed serial interval or estimate generation time.

**Fig. S7.**
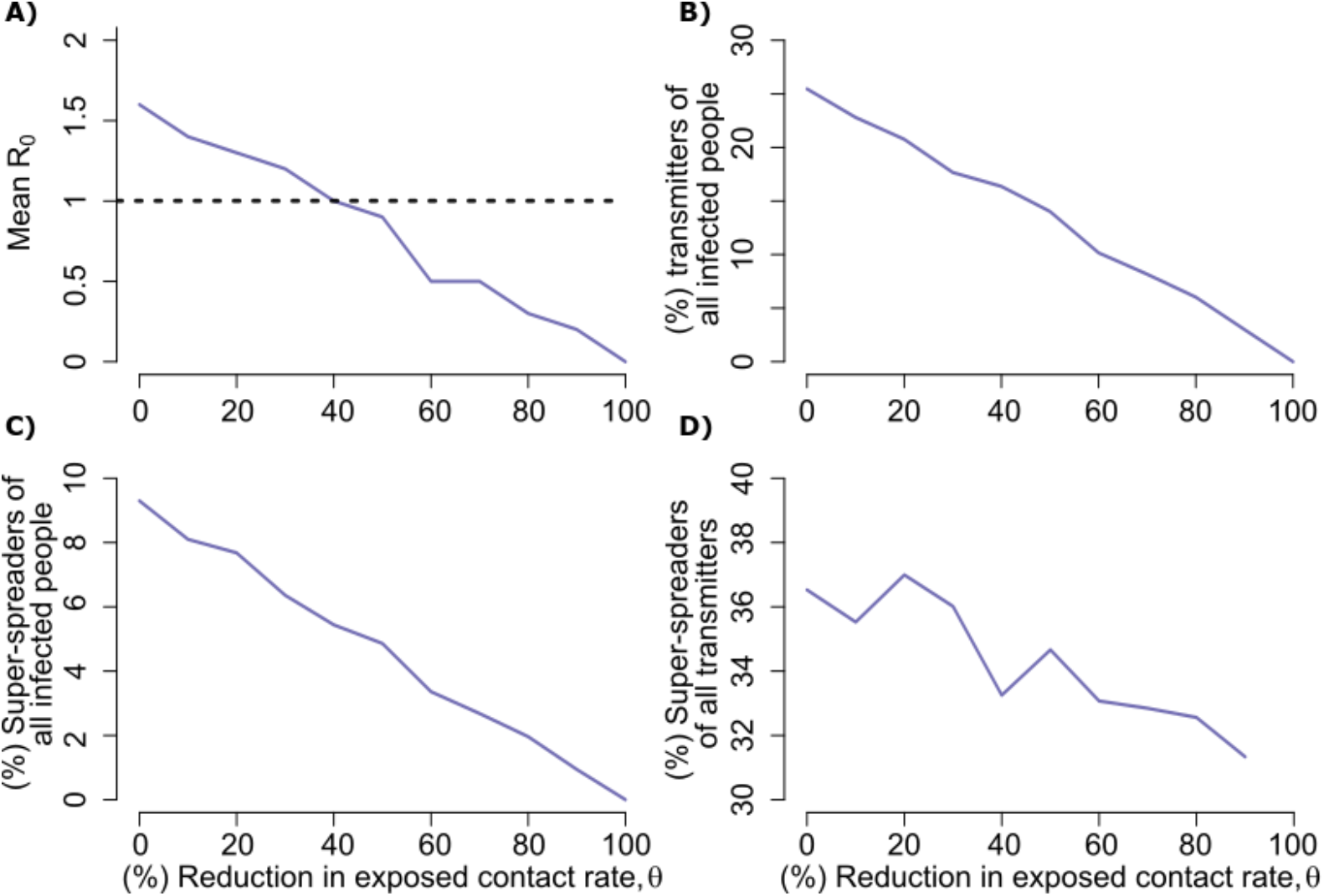
Potential impact of population physical distancing on SARS-Co-V2 epidemiology. **A**. Mean reproductive number **B**. Percent transmitters of all infected people **C**. Percent superspreaders (individual R0>5) of all infected people **D**. Percent super spreaders of all transmitters. Transmitters are defined as infected people who generate at least one secondary infection.

**Fig. S8.**
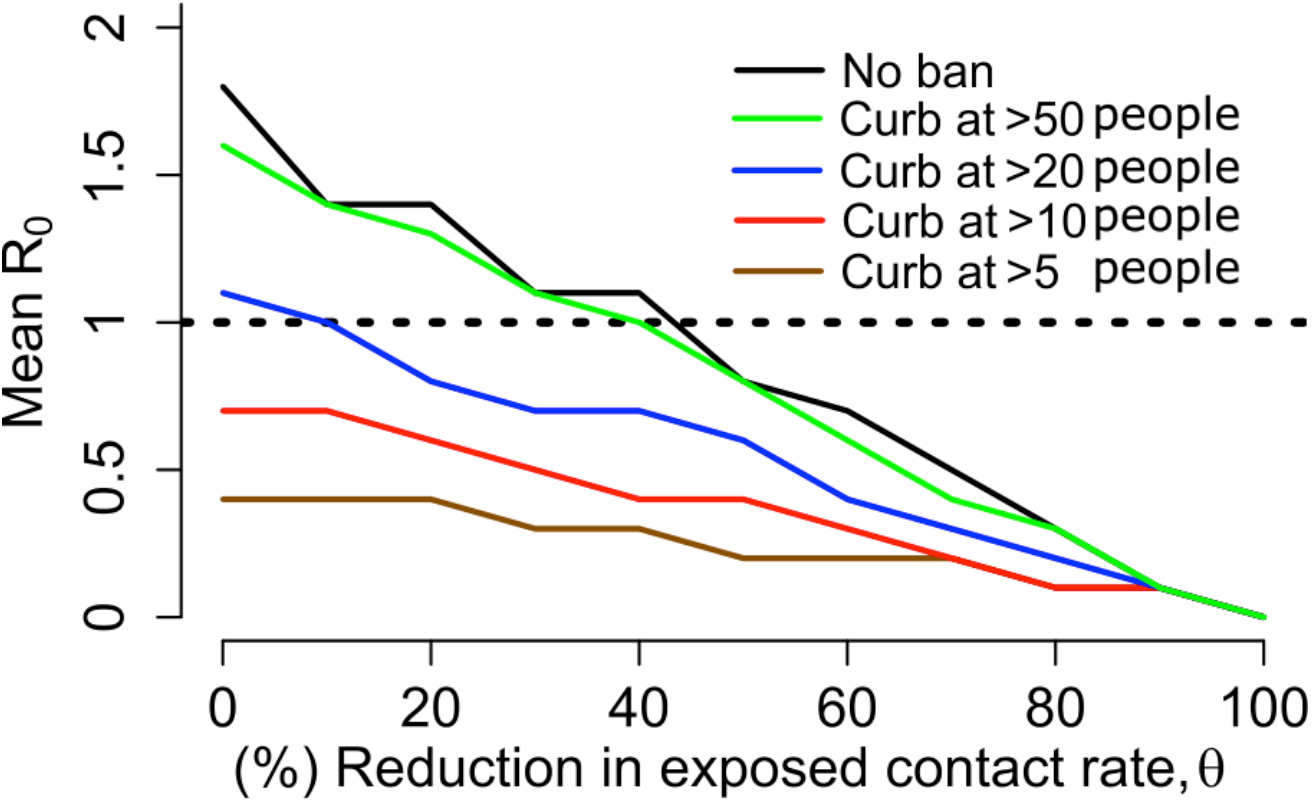
Potential impact of enhanced physical distancing only within high exposure contact networks on SARS-CoV-2 epidemiology. Simulations assume limitation of exposed contacts only among daily exposures of more than 5, 10, 20 or 50 people. Mean reproductive number decreases below one with only marginal decreases in overall rate of exposure contacts when contacts are limited to fewer than 20 people.

**Fig. S9.**
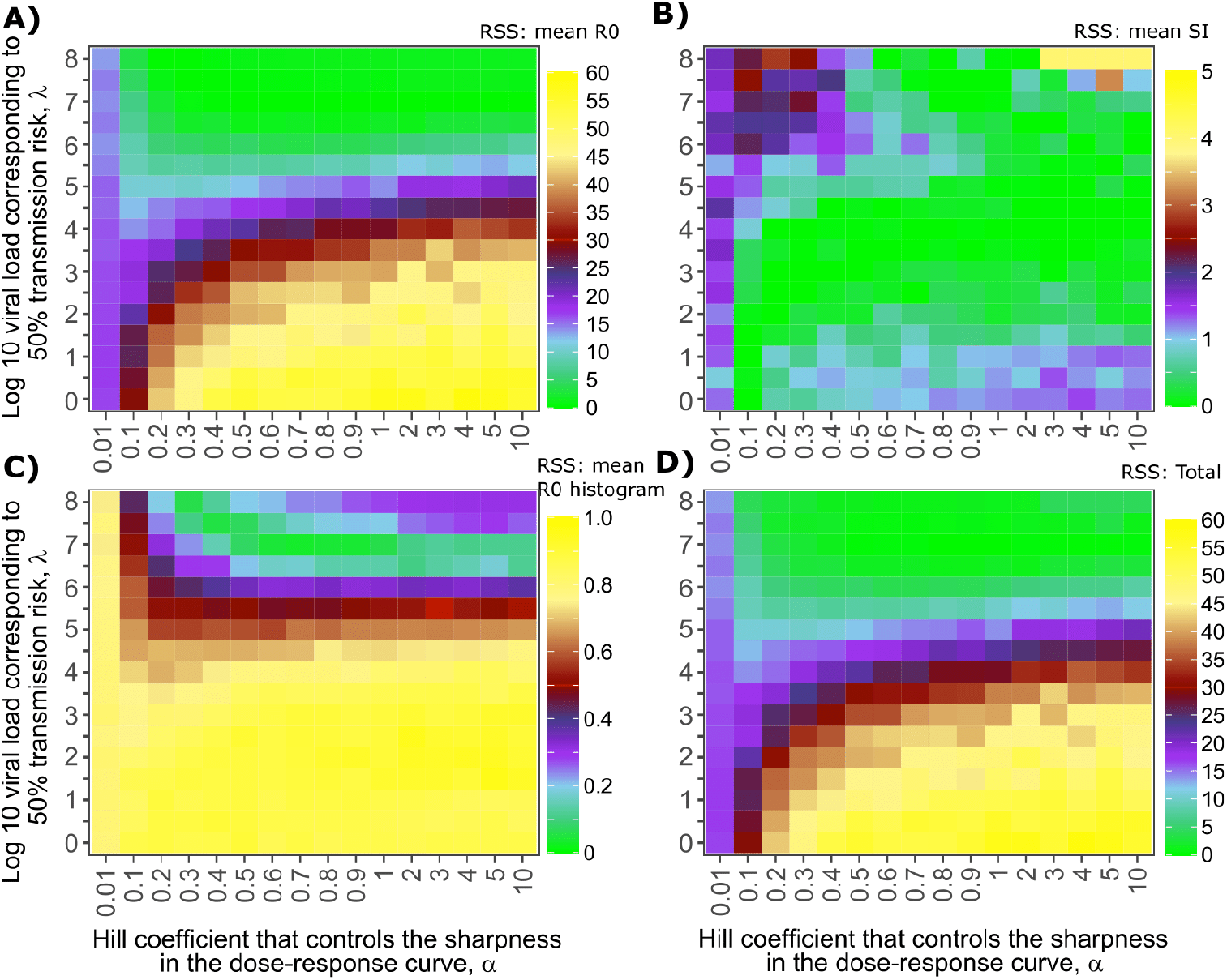
Sensitivity analysis of transmission curve parameter for model fit to SARS-CoV-2 data. Effects of varying transmission curve slope (x-axis) and TD50 for infectiousness (y-axis) on fit to **A**. Mean R0, **B**. Mean serial interval, **C**. Cumulative distribution function of individual R0, and **D**. Sum of Errors in A, B and C.

**Fig. S10.**
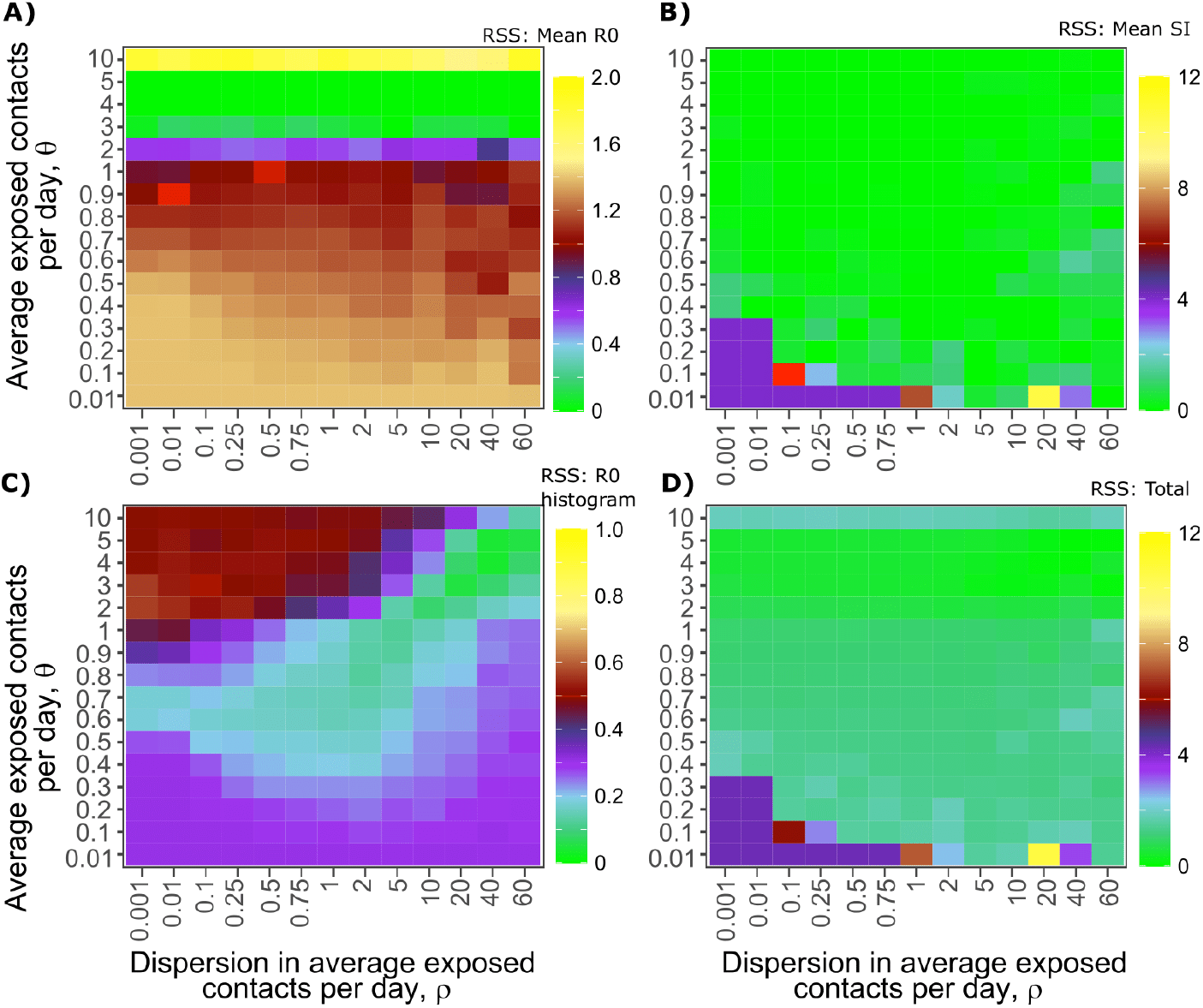
Sensitivity analysis of contact network structure for model fit to SARS-CoV-2 data. Effects of dispersion parameter (x-axis) and average exposed contacts per day (y-axis) on fit to **A**. Mean R0, **B**. Mean serial interval, **C**. Cumulative distribution function of individual R0, and **D**. Sum of Errors in A, B and C.

**Table S1:** Population parameter estimates for simulated SARS-CoV-2 viral shedding dynamics. Parameters are from (doi: https://doi.org/10.1101/2020.04.10.20061325). The top row is the fixed effects (mean) and the bottom row is the standard deviation of the random effects. We also fixed r=10, δE=1/day, q=2.4×10-5/day and c=15/day.

| $\text{Log}_{10}\beta$<br>(virions<sup>-1</sup> day<sup>-1</sup>) | $\delta$<br>(day<sup>-1</sup><br>cells<sup>-k</sup>) | k<br>(-) | $\text{Log}_{10}\pi$<br>(log<sub>10</sub> day<sup>-1</sup>) | m<br>(day<sup>-1</sup><br>cells<sup>-1</sup>) | $\text{Log}_{10}\omega$<br>(day<sup>-1</sup> cells<sup>-1</sup>) |
| --- | --- | --- | --- | --- | --- |
| -7.23 | 3.13 | 0.08 | 2.59 | 3.21 | -4.55 |
| 0.2 | 0.02 | 0.02 | 0.05 | 0.33 | 0.01 |

## References

1 https://coronavirus.jhu.edu/map.html.

2 He, X. et al. Temporal dynamics in viral shedding and transmissibility of COVID-19. Nat Med 26, 672–675, doi:10.1038/s41591-020-0869-5 (2020).

3 Ganyani, T. et al. Estimating the generation interval for coronavirus disease (COVID-19) based on symptom onset data, March 2020. Euro Surveill 25, doi:10.2807/1560-7917.ES.2020.25.17.2000257 (2020).

4 Endo, A., Centre for the Mathematical Modelling of Infectious Diseases COVID-19 Working Group, Abbott, S., Kucharski, A. & Funk, S. Estimating the overdispersion in COVID-19 transmission using outbreak sizes outside China. Wellcome Open Res 5, doi:10.12688/wellcomeopenres.15842.3 (2020).

5 Lloyd-Smith, J. O., Schreiber, S. J., Kopp, P. E. & Getz, W. M. Superspreading and the effect of individual variation on disease emergence. Nature 438, 355–359, doi:10.1038/nature04153 (2005).

6 Bi, Q. et al. Epidemiology and transmission of COVID-19 in 391 cases and 1286 of their close contacts in Shenzhen, China: a retrospective cohort study. Lancet Infect Dis, doi:10.1016/S1473-3099(20)30287-5 (2020).

7 L., H., P., D., I., C. & et al. High SARS-CoV-2 Attack Rate Following Exposure at a Choir Practice — Skagit County, Washington, March 2020.. MMWR Morb Mortal Wkly Rep 69:606-610. (2020).

8 Park, S. Y. et al. Coronavirus Disease Outbreak in Call Center, South Korea. Emerg Infect Dis 26, 1666–1670, doi:10.3201/eid2608.201274 (2020).

9 Cowling, B. J., Fang, V. J., Riley, S., Malik Peiris, J. S. & Leung, G. M. Estimation of the serial interval of influenza. Epidemiology 20, 344–347, doi:10.1097/EDE.0b013e31819d1092 (2009).

10 Brugger, J. & Althaus, C. L. Transmission of and susceptibility to seasonal influenza in Switzerland from 2003 to 2015. Epidemics 30, 100373, doi:10.1016/j.epidem.2019.100373 (2020).

11 Qi, L. et al. Factors associated with the duration of viral shedding in adults with COVID-19 outside of Wuhan, China: a retrospective cohort study. Int J Infect Dis 96, 531–537, doi: 10.1016/j.ijid.2020.05.045 (2020).

12 Cao, B. et al. A Trial of Lopinavir-Ritonavir in Adults Hospitalized with Severe Covid-19. N Engl J Med, doi:10.1056/NEJMoa2001282 (2020).

13 Pawelek, K. A. et al. Modeling within-host dynamics of influenza virus infection including immune responses. PLoS Comput Biol 8, e1002588, doi:10.1371/journal.pcbi.1002588 (2012).

14 Memoli, M. J. et al. Validation of the wild-type influenza A human challenge model H1N1pdMIST: an A(H1N1)pdm09 dose-finding investigational new drug study. Clin Infect Dis 60, 693–702, doi:10.1093/cid/ciu924 (2015).

15 Pebody, R. G. et al. Use of antiviral drugs to reduce household transmission of pandemic (H1N1) 2009, United Kingdom. Emerg Infect Dis 17, 990–999, doi:10.3201/eid/1706.101161 (2011).

16 Goldstein, E. et al. Oseltamivir for treatment and prevention of pandemic influenza A/H1N1 virus infection in households, Milwaukee, 2009. BMC Infect Dis 10, 211, doi: 10.1186/1471-2334-10-211 (2010).

17 Mayer, B. T. et al. Estimating the Risk of Human Herpesvirus 6 and Cytomegalovirus Transmission to Ugandan Infants from Viral Shedding in Saliva by Household Contacts. Viruses 12, doi:10.3390/v12020171 (2020).

18 Boucoiran, I. et al. Nonprimary Maternal Cytomegalovirus Infection After Viral Shedding in Infants. Pediatr Infect Dis J 37, 627–631, doi:10.1097/INF.0000000000001877 (2018).

19 Corey, L. et al. Once-daily valacyclovir to reduce the risk of transmission of genital herpes. N Engl J Med 350, 11–20, doi:10.1056/NEJMoa035144 (2004).

20 Rodger, A. J. et al. Risk of HIV transmission through condomless sex in serodifferent gay couples with the HIV-positive partner taking suppressive antiretroviral therapy (PARTNER): final results of a multicentre, prospective, observational study. Lancet 393, 2428–2438, doi:10.1016/S0140-6736(19)30418-0 (2019).

21 Cohen, M. S. et al. Antiretroviral Therapy for the Prevention of HIV-1 Transmission. N Engl JMed 375, 830–839, doi:10.1056/NEJMoa1600693 (2016).

22 Schiffer, J. T., Johnston, C., Wald, A. & Corey, L. An Early Test-and-Treat Strategy for Severe Acute Respiratory Syndrome Coronavirus 2. Open Forum Infect Dis 7, ofaa232, doi:10.1093/ofid/ofaa232 (2020).

23 Leung, N. H. L. et al. Respiratory virus shedding in exhaled breath and efficacy of face masks. Nat Med 26, 676–680, doi:10.1038/s41591-020-0843-2 (2020).

24 Goyal, A., Cardozo-Ojeda, E. & Schiffer, J. Potency and timing of antiviral therapy as determinants of duration of SARS CoV-2 shedding and intensity of inflammatory response. medRxiv 2020.04.10.20061325, doi:10.1101/2020.04.10.20061325 (2020).

25 Wölfel, R. et al. Virological assessment of hospitalized patients with COVID-2019. Nature 581, 465–469, doi:10.1038/s41586-020-2196-x (2020).

26 Lescure, F. X. et al. Clinical and virological data of the first cases of COVID-19 in Europe: a case series. Lancet Infect Dis 20, 697–706, doi:10.1016/S1473-3099(20)30200-0 (2020).

27 Young, B. E. et al. Epidemiologic Features and Clinical Course of Patients Infected With SARS-CoV-2 in Singapore. JAMA, doi:10.1001/jama.2020.3204 (2020).

28 Kim, J. Y. et al. Viral Load Kinetics of SARS-CoV-2 Infection in First Two Patients in Korea. J Korean Med Sci 35, e86, doi:10.3346/jkms.2020.35.e86 (2020).

29 Brouwer, A. F., Weir, M. H., Eisenberg, M. C., Meza, R. & Eisenberg, J. N. S. Dose-response relationships for environmentally mediated infectious disease transmission models. PLoS Comput Biol 13, e1005481, doi:10.1371/journal.pcbi.1005481 (2017).

30 Lauer, S. A. et al. The Incubation Period of Coronavirus Disease 2019 (COVID-19) From Publicly Reported Confirmed Cases: Estimation and Application. Ann Intern Med 172, 577–582, doi:10.7326/M20-0504 (2020).

31 Du, Z. et al. Serial Interval of COVID-19 among Publicly Reported Confirmed Cases. Emerg Infect Dis 26, 1341–1343, doi:10.3201/eid2606.200357 (2020).

32 World Health Organization. Statement on the meeting of the International Health Regulations (2005) Emergency Committee regarding the outbreak of novel coronavirus (2019-nCoV). (2020).

33 Nishiura, H., Linton, N. M. & Akhmetzhanov, A. R. Serial interval of novel coronavirus (COVID-19) infections. Int J Infect Dis 93, 284–286, doi:10.1016/j.ijid.2020.02.060 (2020).

34 Zhang, Y., Li, Y., Wang, L., Li, M. & Zhou, X. Evaluating Transmission Heterogeneity and Super-Spreading Event of COVID-19 in a Metropolis of China. Int J Environ Res Public Health 17, doi:10.3390/ijerph17103705 (2020).

35 Dillon, A. et al. Clustering and superspreading potential of severe acute respiratory syndrome coronavirus 2 (SARS-CoV-2) infections in Hong Kong. PREPRINT (Version 1) available at Research Square, doi:10.21203/rs.3.rs-29548/v1 (2020).

36 Miller, D. et al. Full genome viral sequences inform patterns of SARS-CoV-2 spread into and within Israel. medRxiv, 2020.2005.2021.20104521, doi:10.1101/2020.05.21.20104521 (2020).

37 van Kampen, J. J. A. et al. Shedding of infectious virus in hospitalized patients with coronavirus disease-2019 (COVID-19): duration and key determinants. medRxiv, 2020.2006.2008.20125310, doi:10.1101/2020.06.08.20125310 (2020).

38 Baccam, P., Beauchemin, C., Macken, C. A., Hayden, F. G. & Perelson, A. S. Kinetics of influenza A virus infection in humans. J Virol 80, 7590–7599, doi:10.1128/JVI.01623-05 (2006).

39 Lessler, J. et al. Outbreak of 2009 pandemic influenza A (H1N1) at a New York City school. N Engl J Med 361, 2628–2636, doi:10.1056/NEJMoa0906089 (2009).

40 Opatowski, L. et al. Transmission characteristics of the 2009 H1N1 influenza pandemic: comparison of 8 Southern hemisphere countries. PLoS Pathog 7, e1002225, doi: 10.1371/journal.ppat.1002225 (2011).

41 Cowling, B. J. et al. The effective reproduction number of pandemic influenza: prospective estimation. Epidemiology 21, 842–846, doi:10.1097/EDE.0b013e3181f20977 (2010).

42 Roberts, M. G. & Nishiura, H. Early estimation of the reproduction number in the presence of imported cases: pandemic influenza H1N1-2009 in New Zealand. PLoS One 6, e17835, doi:10.1371/journal.pone.0017835 (2011).

43 Larremore, D. B. et al. Test sensitivity is secondary to frequency and turnaround time for COVID-19 surveillance. medRxiv, 2020.2006.2022.20136309, doi:10.1101/2020.06.22.20136309 (2020).

44 van Doremalen, N. et al. Aerosol and Surface Stability of SARS-CoV-2 as Compared with SARS-CoV-1. N Engl J Med, doi:10.1056/NEJMc2004973 (2020).

45 Widders, A., Broom, A. & Broom, J. SARS-CoV-2: The viral shedding vs infectivity dilemma. Infect Dis Health, doi:10.1016/j.idh.2020.05.002 (2020).

46 Huang, C.-G. et al. Relative COVID-19 viral persistence and antibody kinetics. medRxiv, 2020.2007.2001.20143917, doi:10.1101/2020.07.01.20143917 (2020).

